# Wearable devices can identify Parkinson’s disease up to 7 years before clinical diagnosis

**DOI:** 10.1101/2022.11.28.22282809

**Authors:** Ann-Kathrin Schalkamp, Kathryn J Peall, Neil A Harrison, Cynthia Sandor

## Abstract

Parkinson’s disease (PD) is a progressive neurodegenerative movement disorder with a latent phase and no currently existing disease-modifying treatments. Reliable predictive biomarkers that could transform efforts to develop neuroprotective treatments remain to be identified. Using UK Biobank, we investigated the predictive value of accelerometry in identifying prodromal PD in the general population and compared this digital biomarker to models based on genetics, lifestyle, blood biochemistry, and prodromal symptoms data. Machine learning models trained using accelerometry data achieved better test performance in distinguishing both clinically diagnosed PD (N = 153) (area under precision recall curve (AUPRC): 0.14± 0.04) and prodromal PD (N = 113) up to seven years pre-diagnosis (AUPRC: 0.07± 0.03) from the general population (N = 33009) than all other modalities tested. Accelerometry is a potentially important, low-cost screening tool for determining people at risk of developing PD and identifying subjects for clinical trials of neuroprotective treatments.

## Introduction

For most patients diagnosed with Parkinson’s disease (PD), 50-70% of nigral dopaminergic neurons will already have degenerated by the time the hallmark motor symptoms manifest and a clinical diagnosis is made (Fearnley & Lees, 1991). Thus, there remains a need to identify cheap, reliable, easily accessible, and sensitive biomarkers to detect early pathological changes, with success in this field likely to be transformative in identifying suitable participants for involvement in clinical trials of potential neuroprotective therapeutics.

It is well recognised that at the point of a clinical diagnosis, multiple prodromal symptoms could have been present for several years including Rapid-Eye-Movement Sleep Behaviour Disorder (RBD), constipation, hyposmia, depression, anxiety, and excessive daytime somnolence with urinary dysfunction, orthostatic hypotension, sub-threshold motor symptoms, and abnormal dopaminergic molecular brain imaging more recently added to the criteria for prodromal PD (Heinzel et al., 2019; Postuma & Berg, 2016, 2019). Multiple previous studies have examined these symptoms, together with fluid, tissue, and imaging biomarkers to determine their sensitivity in identifying prodromal PD (Fereshtehnejad et al., 2017; Fereshtehnejad et al., 2019; Hustad & Aasly, 2020). However, the absence of multi- modal models, which combine the predictive power of multiple data sources, has limited this work (Postuma & Berg, 2019). Furthermore, most studies have tended to compare those with prodromal PD to control cohorts lacking any comorbidity, limiting the translational validity and real-world applicability of these findings. More research is therefore needed to understand the specificity and effective role of prodromal markers in the general population.

Continuous or semi-continuous monitoring for sustained periods would facilitate the identification of clinical change, obtain robust estimates of a subjects’ impairments and capabilities, and detect subtle changes at the earliest possible opportunity. However, such monitoring cannot be achieved through clinical assessments given the limitations of time, cost, accessibility, and sensitivity (Dorsey et al., 2020). By contrast digital sensors passively collect data continuously in real-world settings without added cost or effort (Brognara et al., 2019). Preliminary analyses of such digital sensors collecting acceleration and heart rate data have demonstrated the potential for distinguishing those with a clinical diagnosis of PD from those without, as well as added capabilities of monitoring motor progression, and describing sleeping behaviour (Johansson et al., 2018; Schlachetzki et al., 2017; Shah et al., 2020). However, these quantitative motor measures remain largely understudied (Heinzel et al., 2019) with studies often limited by small sample sizes (Del Din et al., 2019), or restricted to analysis only after clinical PD diagnosis (Mirelman et al., 2016; Williamson et al., 2021). Detecting early movement alterations using digital sensors to identify diseases *before* clinical diagnosis is a largely unexplored field with much potential for application in the general population.

This study uses the large, prospective population-based cohort recruited to the UK Biobank (UKBB) to investigate the specificity and real-world applicability of accelerometry as a prodromal marker for PD. Since 2006, data has been collected for >500,000 individuals aged 40-69 years with ongoing passive follow-up of clinical status through routinely collected clinical data, including primary care records, hospital admissions, and notifications to the death registry (Bycroft et al., 2018). Accelerometry data were collected for a randomly chosen subset of this cohort who were approached via email (N = 103,712, collected 2013-2015) (Doherty et al., 2017). Using these data, we sought to determine whether accelerometry data can serve as a prodromal marker for PD, examining its specificity by comparing data from those receiving a diagnosis of PD or already having a diagnosis of PD to both unaffected controls as well individuals diagnosed with related disorders, namely neurodegenerative disorders, movement disorders, and comorbid clinical disorders. We compared the performance of our accelerometry model to models using data from other modalities, such as genetics, blood, lifestyle, prodromal symptoms and accelerometry to determine the best data sources to identify prodromal PD.

## Results

### The UK Biobank provides a large and increasing cohort of individuals diagnosed with Parkinson’s disease

Clinical diagnoses within the UKBB currently derive from multiple sources including self-reported symptoms and diagnoses, hospital records, death records, and primary care data. However, there is no clinical diagnostic validation and in some areas the data collection within the UKBB is incomplete. To ensure that the PD cohort identified was neither an over- or under-estimation, we compared the prevalence in our cohort to that expected based on 2015 UK population statistics (Parkinson’s UK, 2017).

At baseline data collection, 967 subjects had previously received a diagnosis of PD. An additional 2869 subjects were diagnosed by March 2021 giving a total of ∼0.76% of UKBB participants had been diagnosed with PD at that time point. Annual incidence of PD increased as the cohort aged (Supplementary Figure 1A), with the highest number of diagnoses made amongst those aged 70-80 years (Supplementary Figure 1B). Overall, the observed number of PD cases was slightly lower than that expected based on 2015 UK population statistics (2015 expected: 2252.61, 2015 observed: 1984; Supplementary Figure 1C and Figure 1), with 5255 expected by 2030 assuming no further deaths were to occur. Although we have not been able to review each individual included in this study face-to-face, as is often done in deeply phenotyped, disease-specific cohorts, and the date of diagnosis may lack accuracy due to incomplete coverage of the electronic health records in terms of time and clinics, in general we found that the prevalence and incidence of PD in the UKBB cohort closely resembled that expected from such a population.

**Figure 1:**
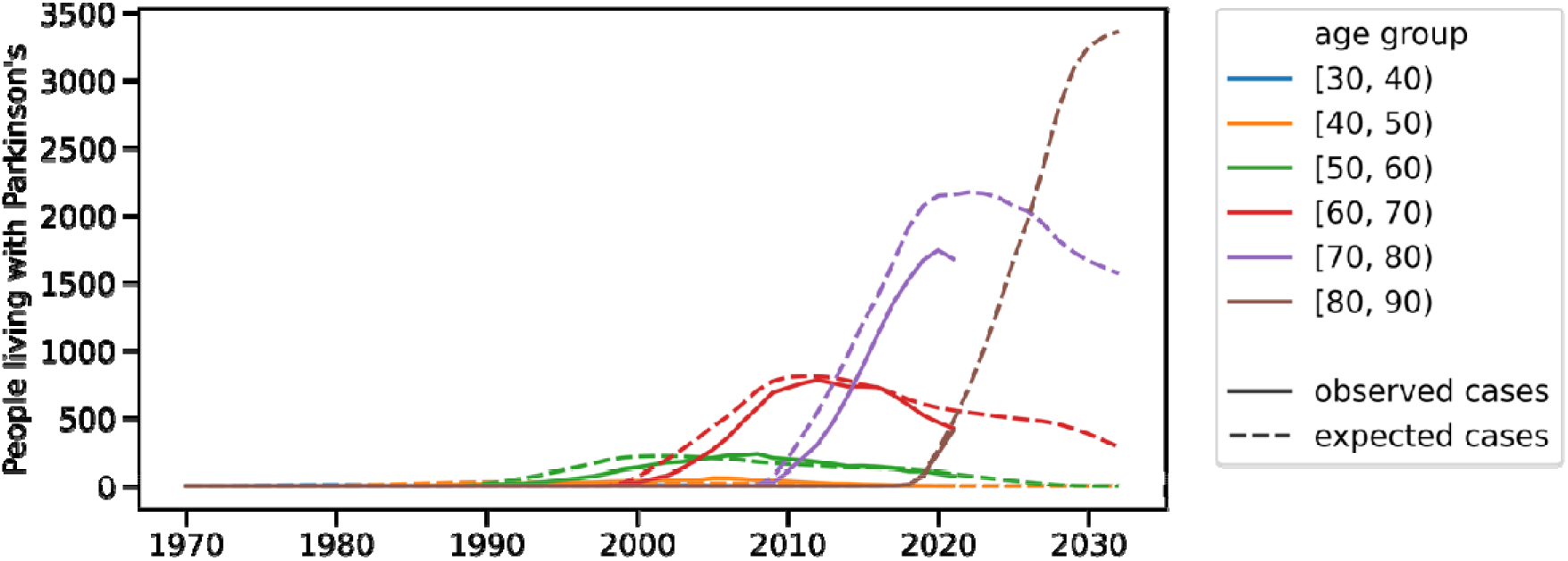
Estimated and Observed Prevalence of Parkinson’s disease in UK Biobank. Estimated (dashed) and observed (solid) number of people living with PD in the UK Biobank over time within age groups is shown. Estimated number of cases uses the population-based UK statistics from 2015 (Parkinson’s UK, 2017)

### Sensor-measured acceleration is reduced several years prior to Parkinson’s disease diagnosis

We compared the average acceleration for each hour of the day over a 7-day period between the diagnostic groups. At the time of or within two years after accelerometry data collection, 273 subjects were diagnosed with PD (mean years since diagnosis: 5.04±6.37; Supplementary Table 1, Figure 2A). An additional 196 individuals received a new PD diagnosis more than two years after accelerometer data collection (mean years to diagnosis: 4.33±1.30, Figure 2A). The prodromal group was significantly older than the diagnosed group (t-statistic = 3.26, d.o.f. = 453, p-value = 1.2×10^-3^, 95% CI = [0.65, 2.61], Cohen’s d = 0.298) and were therefore not directly compared in this initial analysis. To compare the diagnosis groups with an appropriate control group, we randomly sampled age- and sex-matched unaffected controls for each subject diagnosed with PD and each subject that will get a diagnosis of PD. Prodromal (from 7am: p-value = 1.9×10^-4^ to 12am: p-value = 4.5×10^-4^) and diagnosed PD cases (from 7am: p-value = 3.7×10^-5^ to 12am: p-value = 1.4×10^-3^) both showed a significantly smaller acceleration profile over all hours between 7am and 12am than age- and sex-matched unaffected controls (Figure 2B, Supplementary Table 3). No differences in average acceleration during the hours from 12am to 7am were found for prodromal (from 12am: p-value = 0.538 to 7am: p-value = 0.141) or diagnosed cases (from 12am: p-value = 0.586 to 7am: p-value = 0.264) compared to unaffected controls. A reduction in acceleration thus can be observed several years prior to clinical diagnosis.

**Figure 2:**
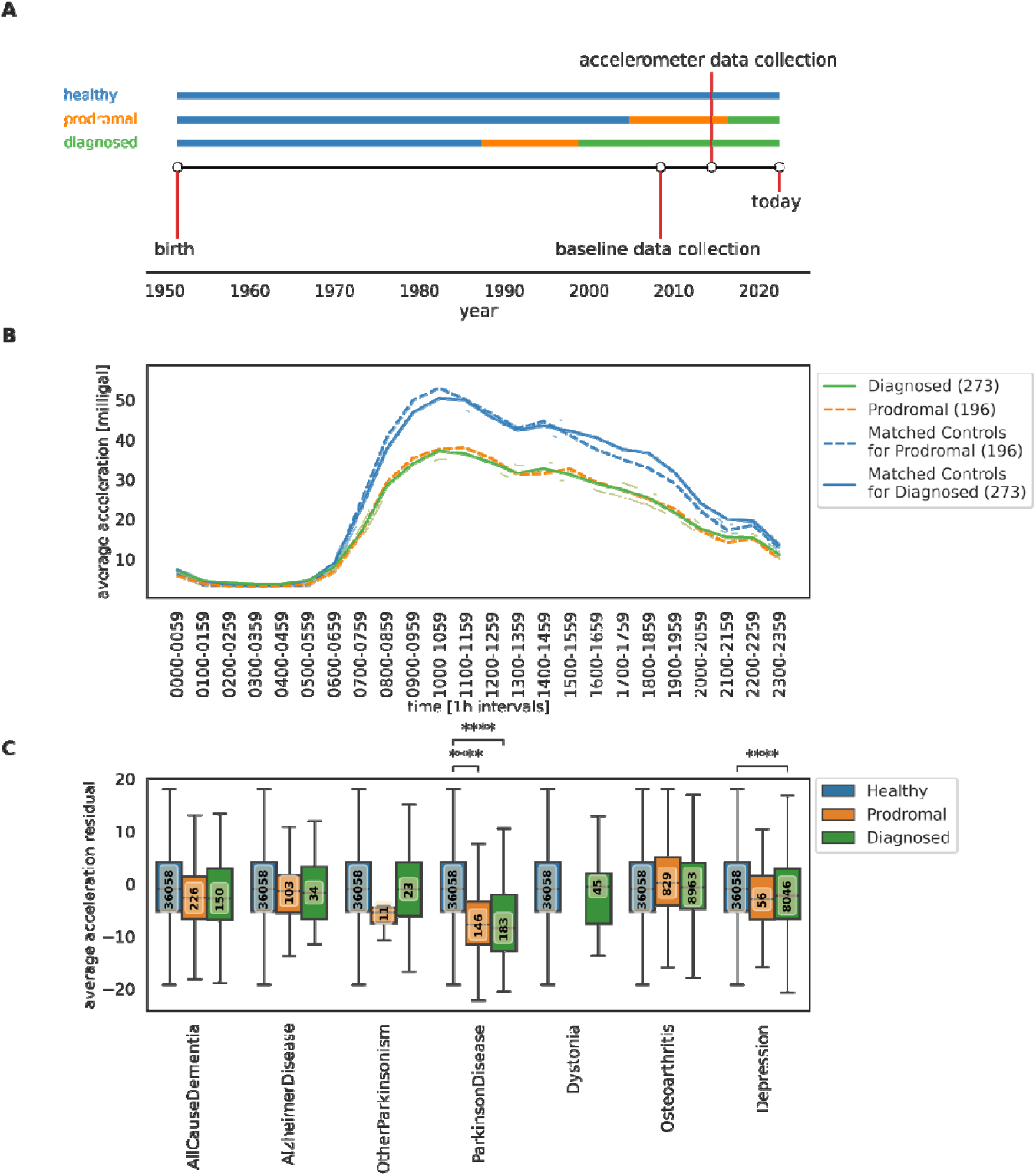
Reduction in acceleration prior to diagnosis is unique to Parkinson’s disease. [A] Baseline data were collected between 2006 and 2010; accelerometry data was gathered for a subset between 2013 and 2015. Diagnosed cases (green) were diagnosed prior to or within the subsequent two years of accelerometry data collection. Prodromal cases (orange) were diagnosed two or more years after accelerometry data collection. [B] Average acceleration in milligal (0.01 mm/s^2^) is shown in one-hour intervals over the course of one day. Group means for prodromal subjects (N = 196, orange, dashed), unaffected controls matched to the prodromal ones (N = 196, blue, dashed), diagnosed subjects (N = 273, green, solid), and unaffected controls matched to the diagnosed ones (N = 273, blue, solid) is plotted with the respective 95% confidence interval. [C] Boxplots for residual (age-, BMI-, and sex-corrected through unaffected control cohort) no-wear time bias corrected average acceleration after removal of cases diagnosed with comorbid depression or PD are shown for seven disease groups and unaffected controls. Analyses without these adjustments can be found in Supplementary Figure 4. For each disease group we differentiate between diagnosed (green), prodromal (orange), and healthy (blue). The number of individuals in each group is indicated in the central box. Significance of group differences (two sided T-test) are indicated with star symbols, where significance is reached with a 0.05 Bonferroni-corrected threshold of 2.38×10^-3^ (ns: 2.38×10^-3^ *<* p *<*= 1, *: 2×10^-4^ *<* p *<*= 2.38×10^-3^, **: 2×10^-5^ *<* p *<*= 2×10^-4^, ***: 2×10^-6^ *<* p *<*= 2×10^-5^, ****: p *<*= 2×10^-6^).

### Reduced sensor-detected acceleration at the prodromal stage is unique to Parkinson’s disease

As physical activity varies between individuals irrespective of health status, we explored whether the observed reduction in acceleration was unique to PD or whether it could also be observed in other clinical disorders, notably other neurodegenerative and/or movement disorders (Supplementary Table 1). We calculated residual average acceleration corrected for age, sex, and BMI via linear regression modes learned on unaffected controls (N = 36058) as these variables showed significant associations with average acceleration and differed between diagnoses groups (Supplementary Table 4). As anticipated, several subjects were diagnosed with multiple comorbidities (Supplementary Figure 2): for example, in the ‘AllCauseDementia’ group, 4.7% had also been diagnosed with PD and 63.5% with depression. In this setting, those with a comorbid PD diagnosis were excluded. As depression is a potential prodromal marker of PD (Heinzel et al., 2019), individuals with a co-diagnosis of depression were also removed.

There was a significant reduction in residual average acceleration in diagnosed PD (t-statistic = 6.25, p-value = 4.17×10^-10^) and prodromal PD (t-statistic = 5.69, p-value = 1.3×10^-8^) compared to unaffected controls (Figure 2C, Supplementary Table 10), but no significant difference between individuals with prodromal and diagnosed PD (p-value = 0.88). That the reduction of acceleration in prodromal PD is consistent with the level observed in the diagnosed PD group, suggests that impaired locomotion is detectable several years prior to a clinical diagnosis of PD. We did not find a significant effect of PD treatment on average acceleration in those diagnosed with PD (Supplementary Methods 1, Supplementary Figure 6). Of those investigated, ‘Depression’ was the only other disorder found to show a reduction in acceleration following diagnosis. None of the disorders investigated were found to have a reduction in acceleration prior to diagnosis that persisted following clinical diagnosis, as was observed for PD (Figure 2C, Supplementary Table 10). In addition, subjects included in the ‘AllCauseParkinsonism‘ group demonstrated the same between-group differences as PD, but this effect was lost when those with a PD diagnosis were excluded (Figure 2C, Supplementary Figure 4). Following removal of the PD cases, the ‘OtherParkinsonism’ group consisted of those diagnosed with secondary parkinsonism or other degenerative disease of the basal ganglia (Supplementary Table 2), for which no group differences were observed, likely due to small sample sizes (prodromal group N = 11, diagnosed group N = 23). Overall, the finding of a reduction in acceleration both prior to and following diagnosis was unique to PD, suggesting this measure to be disease specific with potential for use in early identification of individuals likely to be diagnosed with PD.

### More sleep phenotypes are disrupted in prodromal Parkinson’s disease than in related disorders

We downloaded and extracted sleep features from raw accelerometry data for individuals with any of the discussed disorders and unaffected controls; data from a total of 65901 individuals were processed. We labelled physical activity classes using a pretrained Random Forest (Walmsley et al., 2021). The night-time period was defined as 11pm to 6:59am and daytime as 07am to 10:59pm. As done for average acceleration above, we corrected these extracted features for age, sex, and BMI as learned from the unaffected controls (N = 36058) with linear regression models. Sleep features derived from acceleration data indicated reduced quality and duration of sleep both in the prodromal phase and after having been diagnosed with PD (Figure 3, Supplementary Tables 11-15). Compared to both unaffected controls and the prodromal PD group, individuals with a diagnosis of PD slept for fewer hours overall (controls: p-value = 1.59×10^-10^, prodromal: p-value = 1.78×10^-5^), had fewer consecutive hours of sleep (controls: p-value = 9.66 ×10^-38^, prodromal: p-value = 2.98 ×10^-5^), and slept more frequently during the day (controls: p-value = 1.02×10^-30^, prodromal: p-value = 3.13×10^-5^). Prodromal PD cases (p-value = 5.62×10^-7^) and those diagnosed with PD (p-value = 6.5×10^-4^) woke up more frequently during the night compared to unaffected controls, with no significant difference between prodromal and diagnosed PD cohorts (p-value = 0.22). Individuals with prodromal PD slept longer than unaffected controls (p-value = 1.59×10^-10^) and diagnosed PD cases (p-value = 1.78×10^-5^).

**Figure 3:**
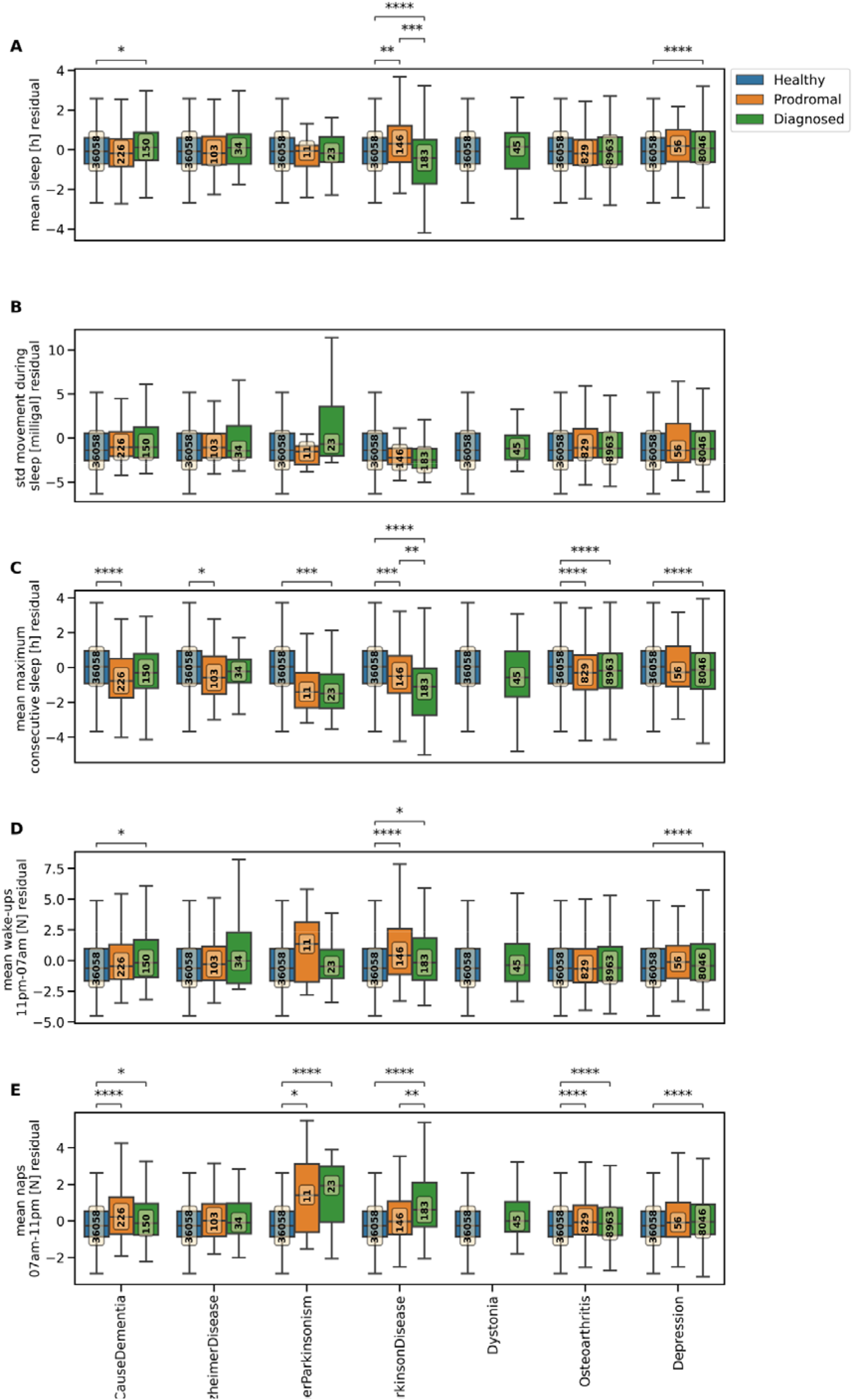
Quality and duration of sleep are reduced in diagnosed but not prodromal Parkinson’s or any other disorder. Boxplots for five residual (age-, BMI-, and sex-corrected through unaffected control cohort) accelerometry derived sleep features after removal of cases diagnosed with comorbid depression or PD are shown for five disease groups and unaffected controls. Supplementary Figure 5 shows the same analyses without exclusion of cases diagnosed with comorbid depression. Due to the covariate correction including a subtraction of the effects of the covariates the residual is displayed and does not reflect the true value of the variable leading to potentially negative values. For each disease group we differentiate between diagnosed (green), prodromal (orange), and healthy (blue). The number of individuals in each group is indicated in the central box. Significance of group differences (two sided T- test) are indicated with star symbols, where significance is reached with a 0.05 Bonferroni-corrected threshold of 2.38×10^-3^ (ns: 2.38×10^-3^ *<* p *<*= 1, *: 2×10^-4^ *<* p *<*= 2.38×10^-3^, **: 2×10^-5^ *<* p *<*= 2×10^-4^,

Examination of the other diagnostic cohorts identified less deterioration in the sleep measures than were found in PD, especially not in the prodromal stage (Figure 3, Supplementary Tables 11-15). Across the distinct diagnostic groups, the length of uninterrupted sleep was the most frequently observed feature to differ between groups. ‘OtherParkinsonism’ and ‘Depression’ showed reductions only after diagnosis whereas ‘AllCauseDementia’ and ‘AlzheimerDisease’ demonstrated reductions only in the prodromal stage. The ‘Osteoarthritis’ cohort was the only one apart from PD in which reductions in consecutive sleep length were observed in both prodromal and diseased stage. Average sleep duration and number of nocturnal awakenings were higher in those in the ‘AllCauseDementia’, ‘Osteoarthritis’, and ‘Depression’ cohorts following diagnosis however, none of the diagnostic groups examined presented a reduction in this feature at the prodromal stage, as was found for PD.

### Accelerometry has the capacity to identify those with prodromal Parkinson’s disease from the general population

We next explored the predictive power of accelerometry data in terms of area under precision recall curve (AUPRC) at an individual level using Lasso logistic regression models with average acceleration, age, and sex as features (Supplementary Table 5) following the TRIPOD reporting guidelines (Collins et al., 2015). The AUPRC was chosen as we have unbalanced datasets such that rare events have to be classified (Davis & Goadrich, 2006). The class imbalance/prevalence will be denoted as N_cases/(N_cases+N_control). Average acceleration distinguished those diagnosed with PD (N = 153) from matched unaffected controls (N = 153) with a mean AUPRC of 0.78±0.06 and could identify prodromal PD (N = 113) from matched unaffected controls (N = 153/113) with the same performance. Identifying diagnosed PD and prodromal PD from non-matched unaffected controls (N = 24987) lead to performances of 0.09±0.05 (prevalence = 0.0061) and 0.09±0.02 (prevalence = 0.0045) respectively. Diagnosed PD and prodromal PD could also be identified from a general population cohort including unaffected controls, and prodromal and diagnosed cases of ‘Osteoarthritis’, ‘Dystonia’, ‘OtherParkinsonism’, and ‘AllCauseDementia’ (N = 33009) using only average acceleration with an AUPRC of 0.05±0.04 (prevalence = 0.0034) and 0.06+0.05 (prevalence = 0.0046) respectively. Adding derived physical activity and sleep features (Supplementary Table 5) increased performance of the models to identify individuals diagnosed with PD from the general population to 0.14±0.04 AUPRC and 0.07±0.03 to identify prodromal PD from the general population. The increase in performance compared to the models only using average acceleration was only significant for the diagnosed PD model (p-value = 0.01), not the prodromal PD model. The most robustly selected feature in all settings was mean acceleration during epochs classified as light physical activity which reduced the risk of having/getting PD the higher it was (Supplementary Figure 16F-21F & 23); meaning that a slowness of movement during normal physical activity can be observed in prodromal and diagnosed PD.

### Accelerometry identifies prodromal Parkinson’s disease more accurately than any other risk or prodromal factor

Several modalities have been explored previously for their value in identifying prodromal PD; however, these were often investigated in isolation and in clinically refined cohorts, rather than the general population. Here, we examined genetics, lifestyle, blood biochemistry, recognised prodromal symptoms for PD, as well as accelerometry (Table 1). We trained Lasso logistic regression models on these different modalities to identify diagnosed (N = 153) or prodromal (N = 113) PD from matched unaffected controls, all unaffected controls (N = 24987), or the general population (N = 33009) (Supplementary Table 9). Of note, the latter group including individuals diagnosed with other neurodegenerative or movement disorders. Models trained on lifestyle, serum biochemical blood markers, recognised prodromal symptoms, or genetic factors showed lower AUPRC scores than models trained on accelerometry features (Figure 4A-C, Supplementary Table 5). We compared each modality-specific model to a no-skill classifier (predictors: intercept) (Supplementary Figure 8 & 9) and noted, that for prodromal PD the prodromal symptoms and accelerometry modality each outperformed this baseline for the matched control setting (Supplementary Figure 9, Supplementary Table 6). When all unaffected controls are included, the blood biochemistry modality additionally outperformed the baseline. In the general population setting every modality except lifestyle outperformed the baseline. For identifying diagnosed PD cases again only the prodromal symptoms and accelerometry modality outperformed the baseline model in the matched setting (Supplementary Figure 8, Supplementary Table 6). When using all unaffected controls, each modality outperformed the baseline. We next compared each single modality model to its respective combined one where the accelerometry modality was added to the predictors. In the diagnosed PD models, the combined models always outperformed the single models for all control settings. For prodromal PD adding the accelerometry modality improved the performance for genetics and lifestyle in the matched setting, every modality except prodromal symptoms when using all unaffected controls, and prodromal symptoms in the general population setting. A model using all available modalities, however, did not outperform the single accelerometry model performance in any setting for either identifying diagnosed or prodromal PD cases, which could be due to each modality capturing different degrees of the same information (Supplementary Figure 24, 25). Overall, the accelerometry modality performed best especially in the general population setting. The prodromal symptoms modality showed good performance in identifying prodromal PD when the controls did not include subjects diagnosed with related disorders.

**Figure 4:**
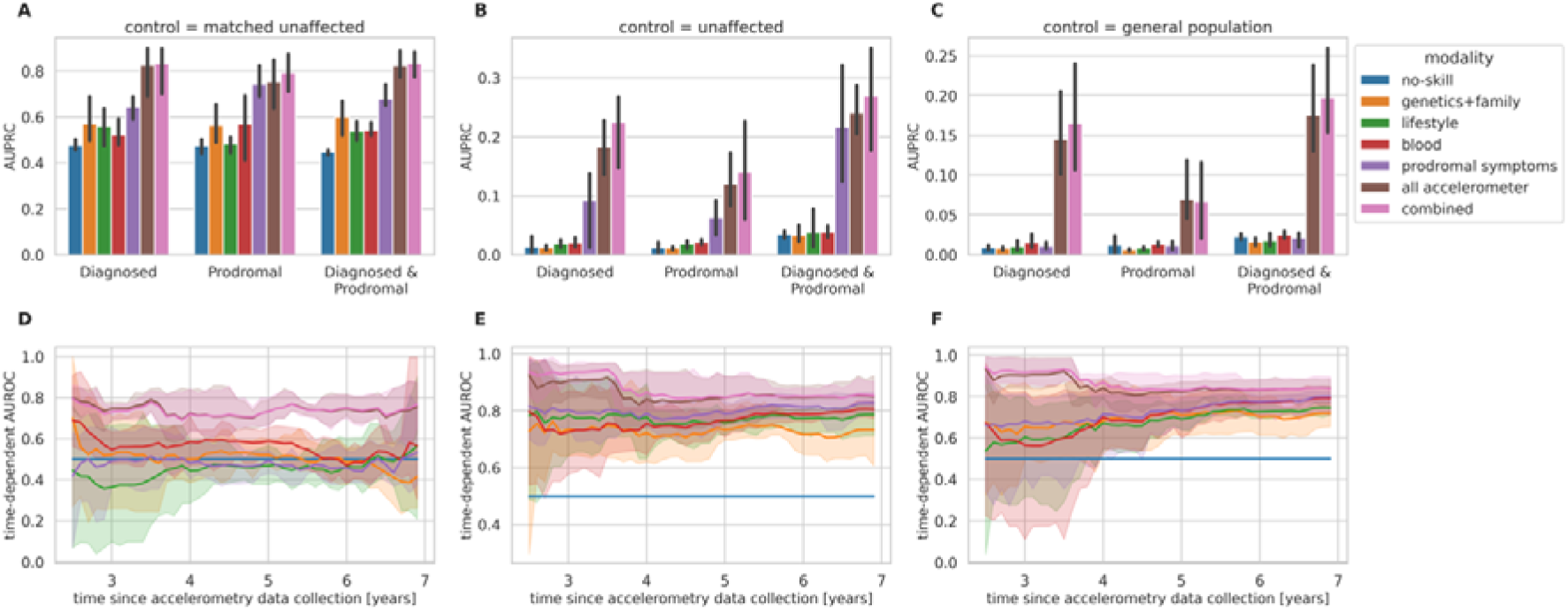
Accelerometry identifies Parkinson’s disease and predicts time to diagnosis better than any other risk factors. [A-C] Bar plots indicate the performance of each logistic regression model using different feature sets (Table 1). The mean area under precision recall curve (AUPRC) are plotted across the five outer cross-validation folds with the error bars indicating the Bonferroni-adjusted 95% confidence interval. We show this for a no-skill, five single modality models, one combined, and one stacked model for three different tasks with two different control groups, [A] matched unaffected controls, [B] all unaffected controls, [C] general population. [D-F] A performance evaluation of the survival models is provided in a time-dependent manner. The time-dependent area under the receiver operator curve (AUROC) of the random survival forests is plotted for several evaluation time-points (years since data collection) together with Bonferroni-adjusted 95% confidence interval for seven years since accelerometry data collection. We show this for [D] a control group made up of matched unaffected controls, [E] a control group including all unaffected controls, and [F] a control group representing the general population.

**Table 1:**
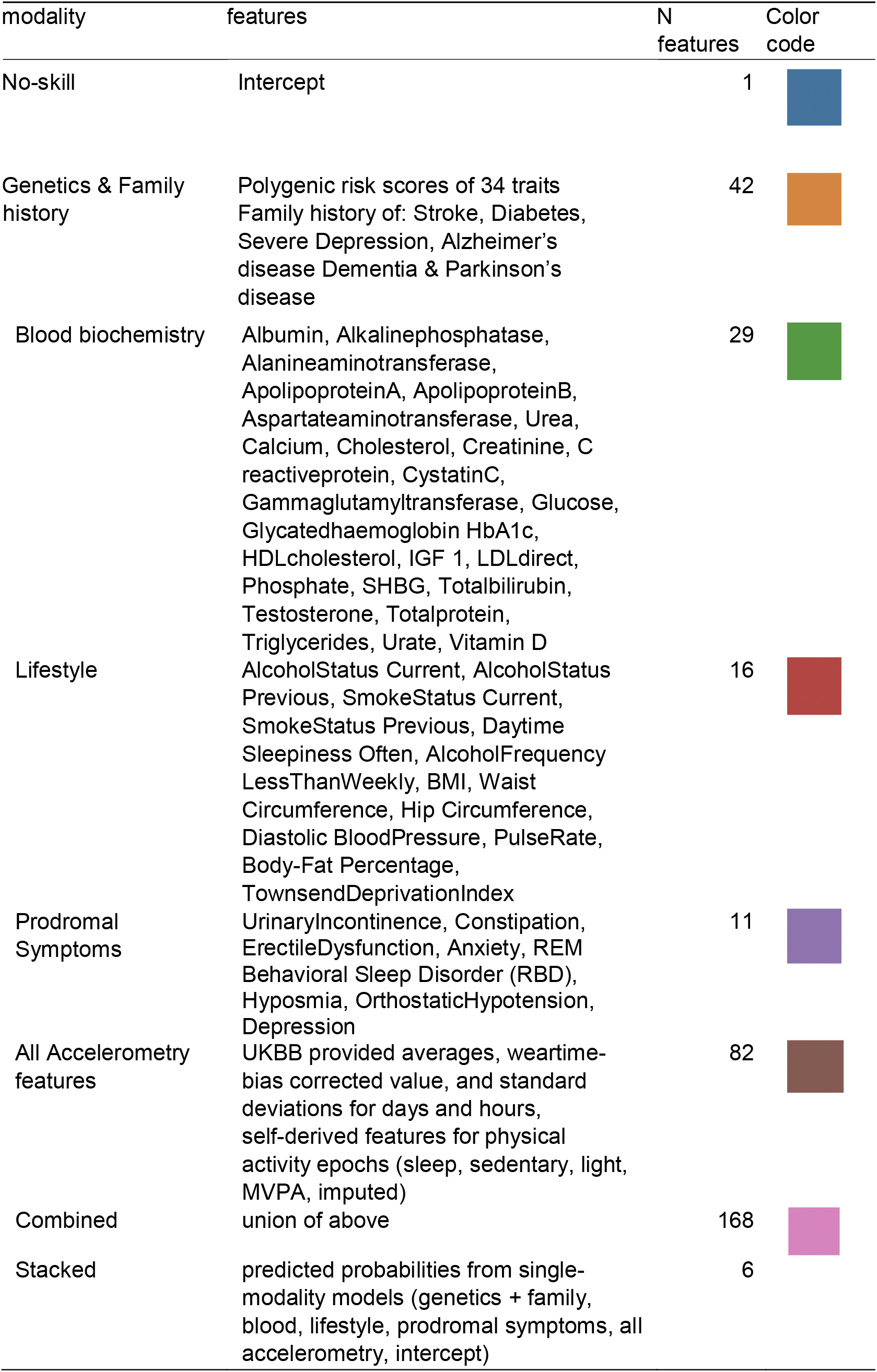
Feature set for each of the modalities.

| modality | features | N<br>features | Color<br>code |
| --- | --- | --- | --- |
| No-skill | Intercept | 1 |  |
| Genetics & Family history | Polygenic risk scores of 34 traits<br>Family history of: Stroke, Diabetes, Severe Depression, Alzheimer's disease Dementia & Parkinson's disease | 42 |  |
| Blood biochemistry | Albumin, Alkalinephosphatase, Alanineaminotransferase, ApolipoproteinA, ApolipoproteinB, Aspartateaminotransferase, Urea, Calcium, Cholesterol, Creatinine, C reactiveprotein, CystatinC, Gammaglutamyltransferase, Glucose, Glycatedhaemoglobin HbA1c, HDLcholesterol, IGF 1, LDLdirect, Phosphate, SHBG, Totalbilirubin, Testosterone, Totalprotein, Triglycerides, Urate, Vitamin D | 29 |  |
| Lifestyle | AlcoholStatus Current, AlcoholStatus Previous, SmokeStatus Current, SmokeStatus Previous, Daytime Sleepiness Often, AlcoholFrequency LessThanWeekly, BMI, Waist Circumference, Hip Circumference, Diastolic BloodPressure, PulseRate, Body-Fat Percentage, TownsendDeprivationIndex | 16 |  |
| Prodromal Symptoms | UrinaryIncontinence, Constipation, ErectileDysfunction, Anxiety, REM Behavioral Sleep Disorder (RBD), Hyposmia, OrthostaticHypotension, Depression | 11 |  |
| All Accelerometry features | UKBB provided averages, wear-time-bias corrected value, and standard deviations for days and hours, self-derived features for physical activity epochs (sleep, sedentary, light, MVPA, imputed) | 82 |  |
| Combined | union of above | 168 |  |
| Stacked | predicted probabilities from single-modality models (genetics + family, blood, lifestyle, prodromal symptoms, all accelerometry, intercept) | 6 |  |

We next evaluated which factors within each modality were considered the most relevant (Supplementary Figure 16-21) and provide a measure of how many features from within a modality were significant in the combined model. We restricted this second analysis to the models build using matched unaffected controls. Features from the accelerometry modality made up the largest portion (67% for the model identifying prodromal PD and 50% for the model identifying diagnosed PD) of the most important features in the combined model (Supplementary Figure 16F, 19F). The second modality in order of importance to the model was genetic markers with the PRS for PD as the found feature, with values of 33% and 25% for prodromal and diagnosed PD respectively. Mean acceleration during epochs that were classified as light activity was the strongest protective factor for every model (Supplementary Figure 16-21). The most important features in the combined model resembled those found in the modality-specific models (Supplementary Figure 16-21). For the models using all unaffected controls, more features were identified as stable across folds, this however decreased when subjects with related disorders were included in the control group. As the accelerometry markers selected as the most relevant also came from the modality with the highest number of features (Table 1), one could argue that their importance was purely due to their dimensionality. We investigated this through a stacked model where the predicted probabilities of the modality-specific models served as input to a final logistic regression model. This performed as well as the combined model (Supplementary Table 7) and also identified the same modalities as the most important ones (Supplementary Figure 22).

### Acceleration-derived phenotypes can predict time to diagnosis

An estimate of time to diagnosis would not only have potential clinical utility but would also be important in clinical trials evaluating the efficacy of disease-modifying or curative therapies. As such, we next explored which modality would be most beneficial in predicting time to clinical diagnosis of PD. A survival random forest model was used to predict time to diagnosis using several features, following the same modality-specific modelling approach described previously for the logistic regression models (Supplementary Table 9) here however we only focus on prodromal PD and modelling their time to diagnosis compared to our three control groups. The model trained on accelerometry features achieved a mean AUROC (from the five-folds time-dependent AUROCs) of 0.74±0.04 when restricted to matched unaffected healthy controls, 0.86±0.06 when prodromal cases are identified from all unaffected controls, and 0.84±0.04 when trained on the general population (Figure 4D-F, Supplementary Table 7). The brier score can be found in Supplementary Figure 28. As survival class (right censored class) outnumbers the amount of people getting a diagnosis of PD within the observed time frame, the AUROC scores should only be evaluated in a comparative manner as the metric can be overoptimistic in this imbalanced setting. The accelerometry model in the could predict the probability of not receiving a PD diagnosis over time significantly better than any other single modality model in all settings (Figure 4D-F, Supplementary Table 8). This finding highlights that acceleration data not only allowed us to predict who would develop PD but also when this diagnosis might be expected.

## Discussion

Here we show the potential of accelerometry as a prodromal biomarker for PD. We found that reduced acceleration manifests many years prior to clinical PD diagnosis. This pre-diagnosis reduction in acceleration was unique to PD and was not observed for any other disorder examined. By comparing the predictive value of accelerometry with other modalities including genetics, lifestyle, blood biochemistry, and prodromal symptoms, we found that no other data type performed better. This improvement in predictive power remained even when assessing our models in a more real-world scenario where the control group contained individuals with related disorders in addition to unaffected controls. Finally, we showed that accelerometry can further predict the time-point at which a PD diagnosis can be expected.

To our knowledge, by using a large sample of individuals who convert to PD *after* data collection, we provide the first demonstration of the clinical value of accelerometry-based biomarkers for prodromal PD in the general population. This work builds on prior clinical data which has demonstrated abnormal motor functioning during the prodromal phase of PD. Darweesh and colleagues demonstrated that impairment in activities of daily living and signs of slowness appeared up to seven years prior to a clinical diagnosis of PD (Darweesh et al., 2017). Similarly, Fereshtehnejad et al. (2019) analysed the temporal evolution of multiple prodromal markers in a longitudinal cohort of patients diagnosed with RBD and highlighted the predictive potential of early motor symptoms to identify prodromal PD up to six years prior to PD diagnosis. However, these works were limited to either a high risk RBD population or time-consuming assessments made in the clinical setting.

Previous work has already explored the use of digital gait markers for diagnosing PD (Del Din et al., 2019; Williamson et al., 2021; Yang et al., 2022). For example, using UKBB data Williamson et al. (2021) demonstrated high accuracy in detecting diagnosed PD cases using free-world collected wrist-worn accelerometer data. However, they focused on prevalent PD cases and did not explore the possibility of using these digital markers to identify PD years *before* clinical diagnosis. To date, the only other study to have used gait-measuring sensors to investigate the prodromal phase of PD was limited to 16 subjects who converted to PD (Del Din et al., 2019). Further, the gait data acquired in that study was collected during a specified task in clinic and not free-living conditions.

Other digital markers have also been investigated for their use as potential prodromal biomarkers, though typically in equally small sample sizes. For example, nocturnal breathing patterns that can be collected at home via radio waves have been shown to predict severity and progression in diagnosed PD cases but were also examined as risk assessment tools for prodromal PD (N = 12) with 75% being labelled as PD before their clinical diagnosis (Yang et al., 2022). Other modalities have been explored considering specificity of the markers by examining other related disorders; for example, cognitive and functional impairment prior to diagnosis have been assessed in several neurodegenerative disorders using UKBB data (Swaddiwudhipong et al., 2022). Transferability to the general population and biomarker specificity have not been reported in previous accelerometry studies due to the data being collected only in PD cohorts; however, the population-based nature of the UKBB data, involving individuals with a broad array of clinical diagnoses, allows us to examine for this potential applicability within the general population. Overall, we identified five major gaps in research that this work aims to address: studying the (1) prodromal phase of PD using passively collected (2) real-world gait-sensor-based data in a (3) large sample size (4) while comparing its performance to other established markers and (5) its generalizability to the general population. We therefore have been able to show, for the first time, the clinical value of accelerometry based biomarkers for prodromal PD compared to other modalities.

As the data is easily accessible and low-cost, using accelerometry data in clinical research and practice is feasible. Currently, electronic health records do not include data from medical-grade wearables. Such data, however, is readily available as smart-devices capable of collecting accelerometer data are used daily by most people (Chandrasekaran et al., 2020). This resource could be linked to electronic health records so that a large retrospective study could be undertaken. A rapidly growing interest in leveraging such data for personalised medicine accelerates this research area with many new digital sensors and wearables being developed (Xu et al., 2022). Challenges to overcome include measurement validity and capability, data privacy, and liability concerns (Simon et al., 2022). Further, processing of the vast amounts of data generated by digital sensors is resource and time intense. As we demonstrated here a single week period of data captured is predictive for several future years, and therefore longer intervals between clinical assessment could be employed, further reducing resource demands. If these limitations are addressed, wearables and other health-sensor devices hold the ability to transition medicine into a digital health era improving accessibility in remote areas, reducing cost, and improving healthcare (Xu et al., 2022).

There are several limitations in this study, the primary one being the lack of replication, although extensive cross-validation was performed to attempt to mitigate against any cohort specific biases. The UKBB provides a unique resource with no equivalent in terms of scale and data volume, collected in such a way as allow for the study of the prodromal phase of multiple disorders retrospectively, and as such no other dataset at present allows for a similar analysis to replicate our findings. For example, the Parkinson’s Progression Marker Initiative (PPMI) cohort although providing longitudinal smart watch data for a prodromal cohort of 158 cases defines prodromal as people at risk, not people who subsequently received a diagnosis of PD, as we have been able to capture in this study. Another dataset, ‘The All of Us’ cohort could provide a valuable future replication resource but at this point is limited by the number of people receiving a diagnosis after data collection. This is likely due, in part, to recruitment being from a wider age range (>18 years in ‘The All of Us’ compared to 40-69 years in the UKBB) with data capture to date being over a shorter period (2018 onwards for ‘The All of Us’ study and 2006 onwards for the UKBB).

Several restraints concerning data availability within the UKBB should be noted. Except for a small subset of individuals, accelerometry data was only collected over one seven-day period. Longitudinal data on acceleration would allow investigation of individual trajectories improving the sensitivity of detection of deterioration in physical activity. Additionally, several clinically recognised PD prodromal markers, such as cerebral dopamine transporter imaging or standardised motor examination scores were not available within the UKBB and therefore, could not be compared to the accelerometry data despite their recognised high predictive power (Brigo et al., 2014). As we chose the time of accelerometry data collection as the defining time point for the group assignment into prodromal and diagnosed, this limits the comparison to other data, like lifestyle and blood, collected prior to this. Further, not all included features were available for all subjects and hence, we trained the models only on a subset of individuals where complete datasets were available, artificially reducing our sample size but allowing greater comparability between models (Table 1). Although capture of accelerometry data in only a subset of the UKBB cohort is a limiting factor of the current study, data availability in the UKBB however, does not reflect the real-world availability of the modalities. For example, genetic data is much more sparsely available in the general population but was prioritised within the UKBB while accelerometry data can be easily and cost-efficiently gathered.

Downloading the raw accelerometry data and the processing pipeline to label the physical activity is time-consuming (30 seconds to download the raw data for one subject ∼250MB, ∼3 minutes to process one subject), thus we processed only the raw data for our identified diagnoses of interest and unaffected controls. This led to the general population missing individuals diagnosed with several other diseases, and thus limiting the transferability of our model to clinical practice as it was not trained on a perfect representation of the general population. This highlights a further limitation being the time-consuming processing involved in analysing the vast amount of data generated by smartwatches if they were to be deployed for screening in the general population. As shown here, a seven-day period of data is capable of predicting events up to seven years into the future with comparably high performance meaning that a continuous assessment is not needed but windows between assessments can span over significant time thus reducing the computational resources needed. A final limitation is our choice of model. Using Lasso logistic regression, we focussed on a highly interpretable model of low complexity. Exploring more advanced models that allow for non-linearity could potentially further increase the performance of the models and hence forms a promising future outlook.

In conclusion, our results suggest that accelerometry collected with wearable devices in the general population could be used to identify those at elevated risk for Parkinson’s disease in an unprecedented scale, and, importantly, these individuals who will likely convert within the next few years can be included in studies for neuroprotective treatments.

## Material and Methods

An overview of the performed analyses and included subjects can be found in Supplementary Figure 3.

### Study Population

The UKBB holds in-depth information on ∼500,000 participants and is approved by the Research Ethics Committee (reference 16/NW/0274). It was accessed under the application code 69610 with data released to Cardiff University. Written informed consent of all participants was obtained by UKBB. We explored PD and related disorders, namely ‘AllCauseDementia’, ‘AllCauseParkinsonism’, ‘AlzheimerDisease’, ‘Dystonia’, ‘Osteoarthritis’, and ‘Depression’. We identified patient groups based on ICD10 and ICD9 codes in the hospital inpatient data (fields 41270 and 41271) and the death registry (fields 40001 and 40002) which were curated from UKBB provided tables and phecodes (Wu et al., 2018), as well as self-reported diagnoses (field 20002). Primary care data was also included (field 42040) using read codes (version 2 and 3) respective to the ICD10 codes as mapped through TRUD NHS Read browser (NHS). The respective codes for each diagnosis can be found in Supplementary Table 2. We distinguished prodromal (incident) and diagnosed (prevalent) cases at the date of accelerometer data collection (field 90003) based on the earliest reported date across all resources and allowed for a two-year margin of error, meaning that patients diagnosed before or within the two years after accelerometer data collection are classified as diagnosed/prevalent cases (Figure 2A) and individuals receiving a diagnosis >2 years later are labelled prodromal/incident cases. Unaffected controls were defined as having no neurological, behavioural disorder or any of the included disorders across all included sources and not having been prescribed Antiparkinsonism drugs by a GP (field 42039) (Supplementary Table 17) or self-reporting (field 20003) usage of Antiparkinsonism drugs (ATC = N04, mapped using Supplementary Table 1 of Wu et al. (2019)). From the set of unaffected controls, we randomly sampled unique age- and sex-matched individuals to our PD patients (1:1). Only participants who passed quality control for the accelerometer data (field 90016) were included. Health-related outcome data is available up to March 2021. Using the same approach, we also identified subjects with recognised prodromal signs and symptoms, namely depression, anxiety, orthostatic hypotension, RBD, hyposmia, urinary incontinence, and constipation. We defined these as prodromal symptoms if they were reported before a PD diagnosis was made.

### Accelerometer data

103,712 participants who agreed to participate after random email recruiting wore an Axivity AX3 wrist-worn triaxial accelerometer on their dominant hand for a 7-day-period. UKBB provides summary statistics (category 1009) describing daily and hourly averages. We augmented the accelerometer data by pre-processing the raw data into time-series data and classifying 30 second intervals into physical activity categories, namely imputed, sleep, sedentary, light, or Moderate to Vigorous Physical Activity (MVPA), with a machine-learning model, using balanced random forests with Markov confusion matrices, using the accelerometer package provided by the Oxford Wearables Group (Willetts et al., 2018). The pre-processing steps we employed are the same as those used to derive the summary statistics from UKBB. These steps include device calibration, resampling to 100 Hz, and removal of noise and gravity (Doherty et al., 2017). From this machine learning labelled time-series data we derived measures of uninterrupted duration, mean movement, and number of interruptions for each physical activity category. For sleep this entailed measures of sleep quality, for example frequency of night-time waking and frequency of daytime napping. For this, we only used complete datasets, so for each subject incomplete hours or days, respective to the measure of interest, were removed. We retained for each measure the highest possible amount of data leading to different data being used for different summary statistics. For example, maximum hours spent continuously in one physical activity class was calculated over all data available whereas the mean hours per 24 hours spent in one activity class was calculated as the mean of only fully covered 24h periods starting at 10am. 10am was chosen as the offset as data collection was implemented to start at that time. This removal of data was performed to avoid biases through higher representation of specific hours of the day. A list of all calculated derived physical activity and sleep phenotypes can be found in Supplementary Table 16.

### Additional data

We merged the accelerometry summary statistics (category 1009), blood biochemistry measures at initial visit (category 17518), physical health measures at initial visit (category 100006), Polygenic Risk Scores (PRS) (category 301), and our derived physical activity phenotypes. We further included age at accelerometer data collection and sex.

### Statistical Analyses

All statistical analyses and model training were carried out in python 3.8 using scipy 1.6.1, pingouin 0.5.1 (Vallat, 2018), scikit-learn 0.23.2 (Pedregosa et al., 2011) and sksurv 0.14.0 (Pölsterl, 2020) packages and figures were generated with seaborn 0.11.1 (Waskom, 2021) and matplotlib. Data retrieval from UKBB was facilitated with an adapted version of the ukbb_parser (Brandes et al., 2020) (https://github.com/aschalkamp/ukbb_parser).

### Prevalence

We validated our established cohort of PD cases by comparing the observed to the expected prevalence. For each year between 1950 and 2021 we identified the number of diagnosed and undiagnosed cases in each age-group. Based on the date of death (field 40007) subjects were excluded from the statistics from their year of death onwards. We calculated the estimated number of PD cases for each year based on the number of people alive in each age group and the prevalence rates for individual age groups from a population-based study from 2015 (Parkinson’s UK, 2017). We extrapolated this expected prevalence until 2030, making the assumption of no deaths taking place.

### Identifying and Adjusting for Covariates

Age and sex are known covariates of acceleration. To address this, we subsampled the unaffected controls in an age- and sex-matched manner. However, prodromal and clinically diagnosed groups differed significantly in age (t-statistic = 3.18, p-value = 1.6×10^-3^). BMI is also a covariate for acceleration (Supplementary Table 4). To address this, we calculated the residuals of average acceleration (field code = 90087) using coefficients for age, BMI and sex learned from the unaffected control group (N = 36082) with a linear regression model including an intercept. This resulted in the removal of some subjects due to missing information of some covariates. When examining the other (non-PD) diagnostic groups, we removed any cases in which comorbid PD was observed to attempt to maintain cohort homogeneity (Supplementary Figure 2). We also removed subjects with a comorbid diagnosis of depression from all other diagnosis groups as this diagnostic group was found to have a significantly reduced acceleration and is a prodromal marker for PD. We compared the residual average acceleration measure between the prodromal and diagnosed groups for each included diagnosis class with two-sided T-tests and Bonferroni-correction and for comparisons including healthy controls with the Welch t-test and Bonferroni-correction as here the sample sizes differ largely between groups. We also computed the residual sleep features, which were age-, BMI- and sex-corrected, using the same method as described above for average acceleration.

### Prediction Models

An overview of the trained models with outcome definition and included features can be found in Supplementary Figure 7. To quantify the predictive power of the acceleration data on an individual level and to compare it to other modalities, we fitted logistic regression models. We identified five modalities: genetics, lifestyle, blood biochemistry, prodromal symptoms, and accelerometry (Table 1). For each modality we included the most recent information available, meaning that for the lifestyle and blood modality we included the features from the initial visit as those were not collected later on, and for the prodromal symptoms we check up until March 2021 (last update of linked clinical records) for the existence of prodromal symptoms preceding a diagnosis of PD. We restricted the dataset to subjects with information available for all five modalities. We estimated the predictive performance of each modality with logistic regression with fitted least absolute shrinkage and selection operator (LASSO) penalty in a nested cross-validation. We chose LASSO to increase sparsity in our model and thus decrease complexity such that the models would be more stable and less prone to overfitting. Logistic regression being one of the simplest algorithms for binary classification tasks was chosen due to its high interpretability and prominence. Three different model types were trained: 1) diagnostic biomarker: identifying diagnosed PD (N = 153) from control, 2) prodromal marker: identifying prodromal PD (N = 113) from control, 3) screening: identifying diagnosed and prodromal PD (N = 266) from control. The control group was either 1:1 sex- and age-matched unaffected controls, all unaffected controls (N = 24987) or a representation of the general population (N = 33009), which included unaffected controls and subjects diagnosed with other disorders such as dementia, dystonia, osteoarthritis, and other forms of parkinsonism (N = 8022, subjects with comorbidities were only included once). We did not include participants with a single diagnosis of Depression into the control group as the presence of depression before diagnosis of PD was included as a predictor.

We trained models on different modalities, always including the covariates age and sex and an intercept: no-skill (only intercept), genetics, lifestyle, blood, prodromal symptoms, all acceleration features, all modalities combined (Table 1). We further train models combining the features of each modality with all accelerometry features. We trained the models in a nested 5-fold cross-validation using a stratified 5-Fold split for both the inner and outer split such that in each fold 20% of the data were used for testing and 80% for the inner fold 20% for validation with grid search for the best Lasso penalty hyperparameter (10 equidistant values between 10^-5^ and 10^4^). The no-skill model using only an intercept, was trained using only outer cross-validation folds as no hyperparameters had to be fitted in the inner fold, as no penalty was applied here. Parameter selection was applied independently to each training fold. Real valued predictors were standardised based on the training data of the outer split to have a standard deviation of one and a mean of zero. Binary data was encoded as 0/1. The sample size (class imbalance), mean, and standard deviation for each included feature for every case and control group are given in Supplementary Table 9. Balanced class weighting was applied to adjust for class imbalances. We report the mean and 95% confidence interval (CI) of the area under the receiver operator curve (AUROC) and the area under the precision and recall curve (AUPRC) on the outer cross-validation splits to compare models (Supplementary Table 5).

Performance of the classifiers was compared using two-sided T-tests with multiple testing accounted for using Bonferroni correction at 0.05. We compared each modality to the no-skill performance and each modality with its single performance and its performance when combined with the accelerometry modality (Supplementary Figure 8 & 9). For the accelerometry model we compare its single performance to that of a model using all modalities. We showed the mean AUROC and AUPRC curves with 1 standard deviation across folds for each model (Supplementary Figure 10 – 15). We determined which disorders were most likely to be misidentified by the models as PD by assessing the mean predicted probability of having PD as assigned by the model on the outer folds of the test data (Supplementary Figure 26). We further investigated feature importance by calculating the mean effect of each predictor over the five outer cross-validation splits. We validated their stability through checking their effect size in each outer cross-validation split. A feature was labelled as important and significant if the mean effect size across folds was significantly different from zero (95% Bonferroni-corrected CI does not cross zero). In addition to the combined model which used the union of the features across all modalities, we further trained a stacked model which took the predicted probabilities from each modality-specific model and integrated it in a final lasso logistic regression model that predicts the overall probability of having or getting a PD diagnosis. Each modality was thus assigned a coefficient with which its prediction contributed to the final prediction. This model was trained on the same outer cross-validation splits as the other models using their predictions on the training data to train the final model and the respective test data for testing. No penalty hyperparameter was fitted here, so no inner cross-validation was performed. The performance was evaluated in a similar fashion with AUROC and AUPRC across folds and the assigned coefficients across folds were evaluated for their stability across folds by examining the mean and standard deviation (Supplementary Table 5).

### Survival Models

We explored the value of each modality to predict the time to diagnosis for the prodromal PD cohort. We did so by first calculating the Pearson correlation (scipy.stats.pearsonr with two- sided) of the residual average acceleration (age- and sex-corrected) and time to diagnosis in the prodromal PD cohort. A simple linear association was not found between residual average acceleration (age- and sex- corrected) and time to diagnosis for the prodromal cases (r = 0.11, p-value = 0.13), i.e. average acceleration did not appear to decline further closer to the date of diagnosis. We then used survival modelling on the prodromal (N = 113) and controls (matched: N = 113, unaffected controls: N = 24987, or population: N = 33009) to predict when each individual would be diagnosed (Supplementary Figure 27). To this end, we used survival random forests with a five-fold stratified cross-validation. The survival random forest is made up of 1000 trees which requires at least 10 samples for a split and 15 samples per leaf. The controls were modelled as right censored, as we did not know whether or when they would receive a diagnosis. We modelled the time from accelerometer data collection to PD diagnosis, thus defining the time of accelerometer collection as time 0. For the prodromal symptoms modality, we hence restricted the time of diagnosis of prodromal symptoms to the date of accelerometer data collection and removed all subsequent diagnoses of prodromal symptoms. The sample size (class imbalance), mean, and standard deviation for each included feature for every case and control group are given in Supplementary Table 9. We reported the time-dependent AUROC (Figure 4D-F) and brier score (Supplementary Figure 28) on the five cross-validation test sets. These metrics are calculated by defining the cases and controls dynamically for several time points with controls transitioning to cases at the time of their PD diagnosis. At each time point based on the current case and control assignment, the predicted case/control assignment is assessed with a standard AUROC using the true positive (sensitivity) and false positive rate (1 – specificity).

## Supporting information

All Supplemental Tables 1-17

## Data Availability

Data from the UK Biobank (ukbiobank.ac.uk/) are available on application to the UK Biobank.

## Data Availability

Data from the UK Biobank (ukbiobank.ac.uk/) are available on application to the UK Biobank

## Code Availability

Code that supports the findings of this study will be made available on GitHub https://github.com/aschalkamp/UKBBprodromalPD.

## Funding

**A.-K. Schalkamp** is supported by a PhD studentship funded by Health and Care Research Wales.

**C. Sandor** is supported by the UK Dementia Research Institute (UK DRI) funded by the Medical Research Council (MRC), Alzheimer’s Society and Alzheimer’s Research UK (AR- UK) and by the Ser Cymru II programme which is part-funded by Cardiff University and the European Regional Development Fund through the Welsh Government.

**N. Harrison** has nothing to declare.

**K. Peall** is funded by an MRC Clinician-Scientist Fellowship (MR/P008593/1).

## Acknowledgements

The work described here is part-funded by the European Regional Development Fund, administered through the Welsh Government. We are also grateful for the Advanced Research Computing at Cardiff. We are also grateful for the valuable comments of Caleb Webber and the input on survival modelling from Valentina Escott-Price.

## Author Contribution

A-K.S. and C.S. participated in designing the study, topic definition, and review of relevant studies. Machine learning models and statistical analyses were designed and implemented by A-K.S.. Figures and tables were done by A-K.S. with the support of C.S.. A-K.S. wrote the first draft. A-K.S., C.S., N.A.H., and K.J.P. contributed to subsequent versions of the manuscript. All authors critically reviewed the paper, all authors have a clear understanding of the content, results, and conclusions of the study and agree to submit this manuscript for publication. The corresponding author (C.S.) declares that all authors listed meet the authorship criteria and that no other authors involved in this study are omitted. C.S. is ultimately responsible for this article.

## Competing interests

The authors declare no competing interests.

## Notes

### Competing Interest Statement

The authors have declared no competing interest.

### Funding Statement

This study was supported by UK Dementia Research Institute (UK DRI) funded by the Medical Research Council (MRC), Alzheimer's Society and Alzheimer's Research UK (AR-UK) and by the Ser Cymru II programme which is part-funded by Cardiff University and the European Regional Development Fund through the Welsh Government. Funding was also received from Health and Care Research Wales. We are also grateful for the Advanced Research Computing at Cardiff.

### Author Declarations

This study used data made available upon application to ukbiobank.ac.uk/.

### Summary of Updates

Major revision has been done to clarify the main message of the paper. Analyses have been repeated on bigger sample sizes and additional analyses have been added.

