## Supplemental Methods, Figures, and Legends for "Wearable devices can identify Parkinson’s disease up to 7 years before clinical diagnosis"

### Supplements

#### Methods & Findings:

1. Medication effect on Parkinson's disease cases

The UKBB captures data on medication usage through its Primary Care dataset and through self-report at the clinical assessments. First, we extracted self-report information (field code = 20003) about usage of Antiparkinsonism drugs (ATC = N04) by using this ATC codec and mapping it to the UK Biobank codes (Wu et al., 2019). 849 UKBB subjects reported taking medication during their initial assessment with 834 belonging to our identified PD cases. These PD cases had a mean time since diagnosis of 4.74 years at the initial assessments. However, this information, could not be used to check whether medication was taken at the time of accelerometry data collection and was hence disregarded for the final analysis.

Thus, we used the primary care prescription records (field code = 42039) to identify participants that i) were ever prescribed medication typically used in the treatment of Parkinson's and parkinsonism AND ii) had received a prescription for it within 10 weeks before data collection, and hence were likely to be medicated during data collection. Read-codes were taken from the UK Biobank documentation (https://biobank.ndph.ox.ac.uk/showcase/refer.cgi?id=594: Parkinson's disease – P2 (all codes referring to medication usage)). A total of 513 UKBB participants have ever been prescribed PD medication. 302 of these are not in the 'AllCauseParkinsonism' group, 206 are among our identified PD cases. 5.37% of the 3837 PD cases in UKBB have been prescribed PD medication at least once. GP information was missing for 2045 identified PD cases which is 53%. If we were to remove those subjects with no records, 11.49% of the PD cases would be considered treated. Comparing the self-report cases with the ones identified with GP records: 440 of the GP record cases were not identified using self-reported data. Conversely, 243 subjects reported PD medication usage at the initial assessment were not reflected in primary care records, perhaps due to incomplete GP record coverage.

We matched the GP records to the accelerometry data using the date of issue and date of accelerometry collection information. Of the PD cases who have accelerometry data available 20 were ever prescribed medication. 19 of those are diagnosed PD and 1 prodromal PD at accelerometry data collection. 6 of these, including the prodromal case, were prescribed medication only after accelerometry data collection (mean days from accelerometry collection to prescription: 464.42±257.28) and were hence assumed to be not medicated during data collection. 14 of the PD cases taking medication were prescribed medication before accelerometry data collection (mean days since prescription to data collection: 104.56±333.1) and of these 13 subjects had not more than 10 weeks between prescription and data collection (mean days from medication prescription to accelerometry data collection: 15.57±9.85). We compared treated (N = 13) and untreated (N = 260) diagnosed PD cases in terms of average acceleration and found no significant differences, potentially due to small sample sizes. We repeated this for residual average acceleration corrected for age, sex, and BMI which lead to the same results (treated N = 10, untreated N = 219). We also repeated this analysis removing subjects who had no GP prescription information available and found the same results.

#### Supplemental Table 1: Overview of study population

For each diagnosis we show summary statistics for the diseased and prodromal groups and their respective unaffected control matches as well as the whole cohort of unaffected controls. We report the mean and standard deviation of the average acceleration (no wear-time bias corrected, field 90087), age at accelerometer data collection, age at baseline data collection, and time to PD diagnosis. We also show the percent of male, depressed and PD cases in each group and the total size of the group.

#### Supplemental Table 2: Codes for the diseases

For each disease and prodromal symptom, we show the associated ICD10, ICD9, read codes and the code used in UKBB for self-report. The ICD9 and ICD10 codes were extracted with phecodes (Wu et al., 2018) and if available the UKBB curated list of codes. The read codes were mapped to the ICD10 codes using the TRUD NHS Read browser.

#### Supplemental Table 3: Differences in acceleration for every hour of the day

We report the statistics of the two-sided T-tests for the comparison of every hour of the day between prodromal Parkinson's disease (PD) cases and their age- and sex-matched unaffected controls as well as diagnosed PD cases and their matched controls. Significance at 0.05 with Bonferroni correction is reached for p-values smaller than $5.2 \times 10^{-3}$.

#### Supplemental Table 4: Significant association of covariates with average acceleration

For the covariates we report the association with average acceleration ('No_wear_time_bias_adjusted_average_acceleration', field 90087). For the real valued covariates, age at accelerometer data collection and body mass index (BMI), we report the statistics for the pearson correlation and for the binary covariate, male sex, we report the statistics for the Mann-Whitney U.

#### Supplemental Table 5: Performance metrics for each of the models

For each model we show the mean and standard deviation on the test sets of the five outer cross-validation folds. The AUROC and the AUPRC are shown.

#### Supplemental Table 6: Significant differences in performance

For each modality-specific model and the combined models, we show the statistics of the two-sided T-tests of the area under precision recall curve (AUPRC) across the five outer folds of the nested cross-validation.

#### Supplemental Table 7: Performance metrics for each survival model

For each modality-specific survival model and the combined model, we show the mean, standard deviation, and standard error of the time-dependent area under receiver operator curve (AUROC).

#### Supplemental Table 8: Significant differences in performance for the survival models

For each modality-specific model and the combined models, we show the statistics of the two-sided T-tests of the mean area under receiver operator curve (AUROC) across the five outer folds of the nested cross-validation.

#### Supplemental Table 9: Features for the prediction models

For each group (prodromal Parkinson's disease (PD), diagnosed PD, unaffected control matched for prodromal PD, unaffected control matched for diagnosed PD, all unaffected controls, population) we report the mean, standard deviation, and sample size for each predictor included in the logistic regression models (prodromal symptoms beforePD) and the survival models (prodromal symptoms beforeacc).

#### Supplemental Table 10: Group comparison for average acceleration

We report the two-sided T-test statistics of the group comparisons for residual (age-, BMI-, and sex-corrected through unaffected control cohort) no-wear time bias corrected average acceleration after removal of cases diagnosed with comorbid depression or PD.

#### Supplemental Table 11: Group comparison for consecutive sleep

We report the two-sided T-test statistics of the group comparisons for residual (age-, BMI-, and sex-corrected through unaffected control cohort) mean maximum consecutive sleep [hours] after removal of cases diagnosed with comorbid depression or PD.

#### Supplemental Table 12: Group comparison for daytime sleeping

We report the two-sided T-test statistics of the group comparisons for residual (age-, BMI-, and sex-corrected through unaffected control cohort) mean number of naps during daytime (7am-11pm) after removal of cases diagnosed with comorbid depression or PD.

#### Supplemental Table 13: Group comparison for sleep duration

We report the two-sided T-test statistics of the group comparisons for residual (age-, BMI-, and sex-corrected through unaffected control cohort) mean sleep duration [hours] after removal of cases diagnosed with comorbid depression or PD.

#### Supplemental Table 14: Group comparison for night-time waking

We report the two-sided T-test statistics of the group comparisons for residual (age-, BMI-, and sex-corrected through unaffected control cohort) mean number of wakeups during night-time (11pm-7am) after removal of cases diagnosed with comorbid depression or PD.

#### Supplemental Table 15: Group comparison for variability of acceleration during sleep

We report the two-sided T-test statistics of the group comparisons for residual (age-, BMI-, and sex-corrected through unaffected control cohort) standard deviation of acceleration during epochs classified as sleep after removal of cases diagnosed with comorbid depression or PD.

#### Supplemental Table 16: Description of derived physical activity features

A list of the calculated physical activity features included in the accelerometry modality additional to the features obtained from UKBB. A description for each is provided alongside an indicator to which activity class the feature belongs.

#### Supplemental Table 17: List of read codes for Anti-Parkinsonism drugs

A list of the read-codes used to identify individuals who were prescribed antiparkinsonism drugs at any timepoint. This list was extracted from the panel 2 UKBB provided algorithmically defined outcomes.

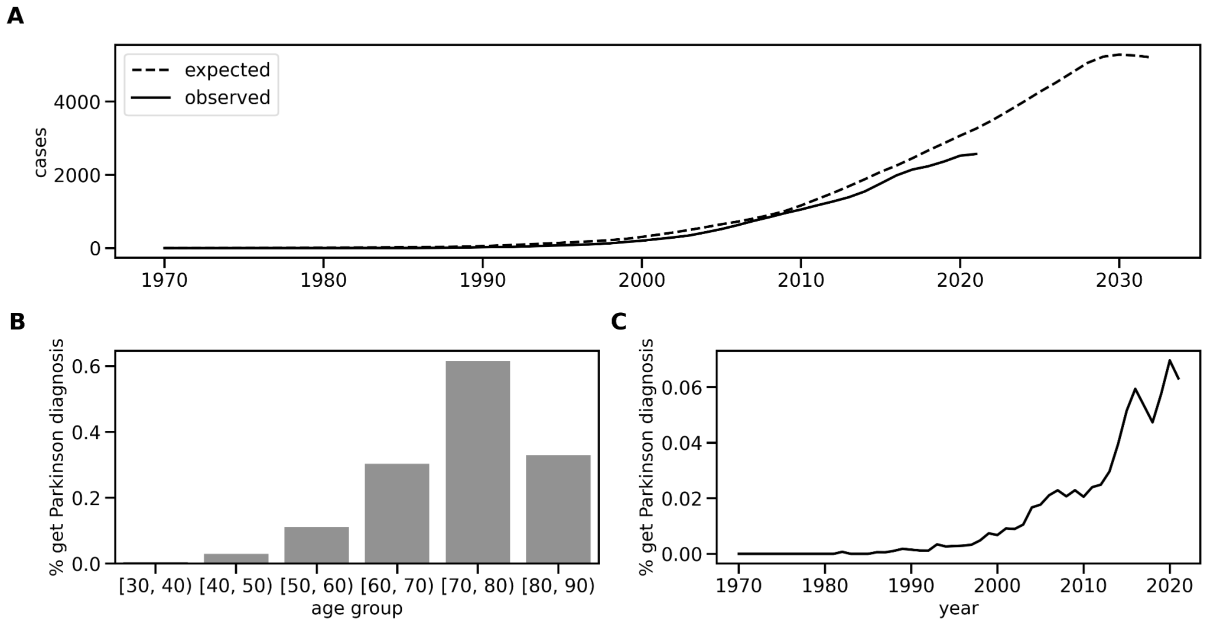


**Supplemental Figure 1: Incidence and prevalence of Parkinson's disease in UK Biobank**.

The plots show (A) the cumulative number of Parkinson's disease (PD) diagnoses expected and observed each year for the UK Biobank, (B) the percentage of people getting a diagnosis within a specific age-range, and (C) the proportion of people getting a new PD diagnosis each year.

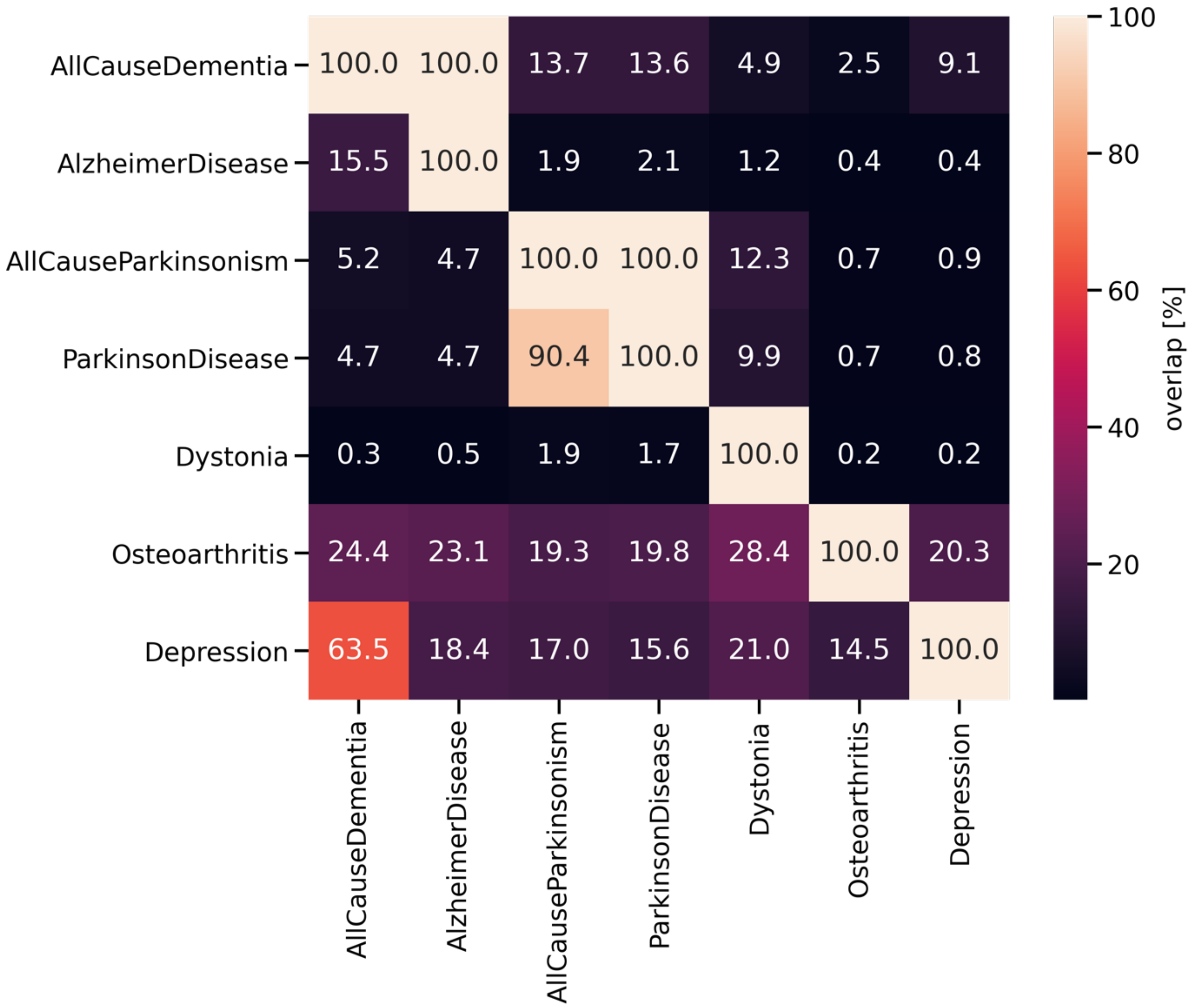


**Supplemental Figure 2: Comorbidities in the UK Biobank.**

Each column indicates the percentage of overlap of a disorder with the other disorders. For example, of all ‘AllCauseDementia’ subjects 63.5% also have a diagnosis of depression and 4.7% also a diagnosis of PD.

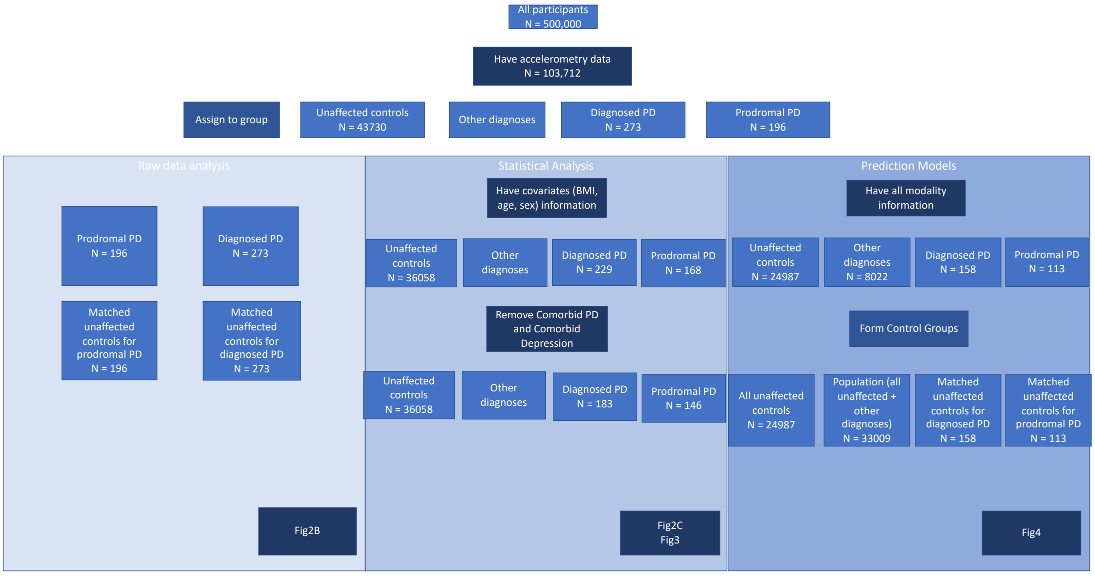


#### Supplemental Figure 3: Subject Flowchart

We show for each analysis how many subjects were included and why they were removed. Starting with the complete UK Biobank dataset, we first focus on those with accelerometry data available. Those are assigned to groups based on our diagnosis extraction method. Three different analyses follow. The first one being done on raw data where unaffected controls are matched to each prodromal and diagnosed case. The second one encompasses statistical analyses for group comparisons, where first only subjects with information on covariates were kept such that residuals were computed, and then comorbid depression and PD were removed. The third analysis trains the prediction models where only subject with complete information on all predictors are kept.

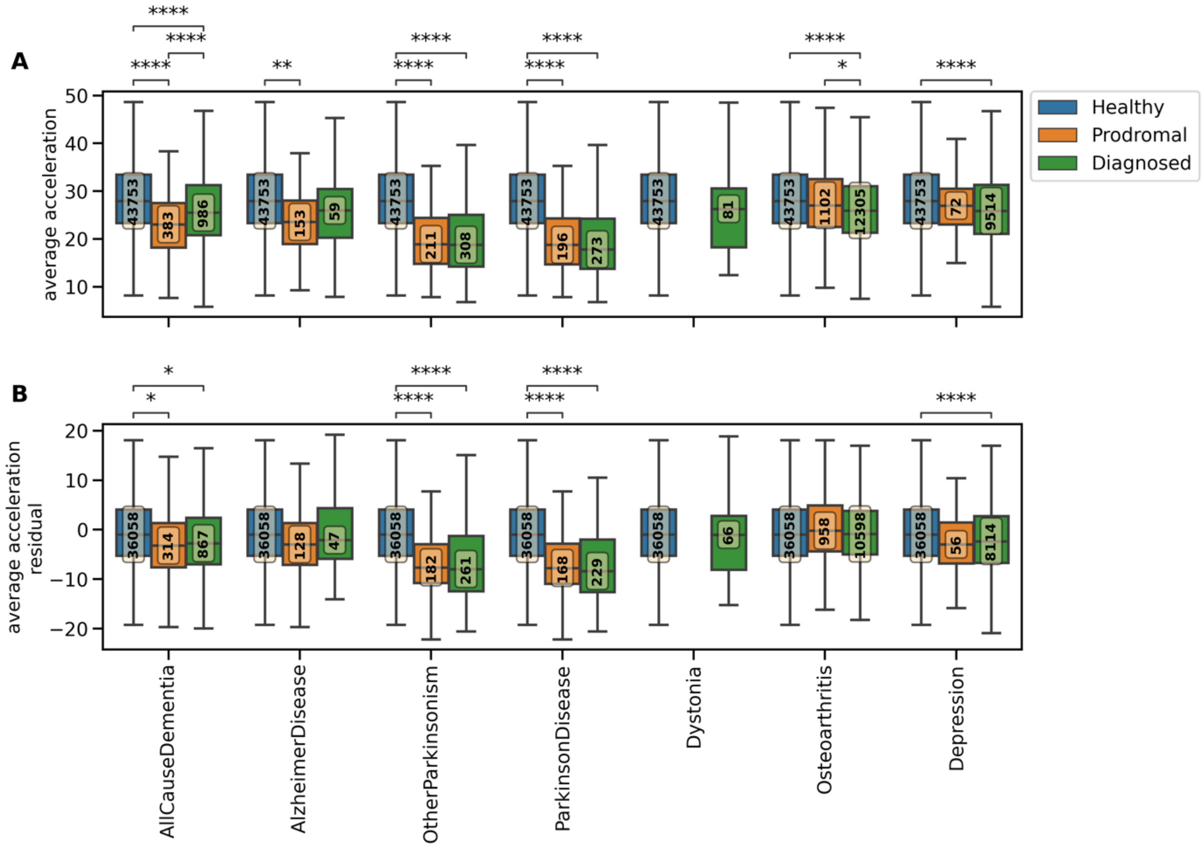


#### Supplemental Figure 4: Reduced acceleration across diseases

Boxplots for no wear-time bias adjusted average acceleration [A] before and [B] after correction for covariates are shown for seven disease groups and unaffected controls. For each disease group we differentiate between diagnosed (green), prodromal (orange), and unaffected (blue). The correction for covariates affects the number of subjects in each group (number in yellow box) as subjects with incomplete information about covariates were excluded. The correction includes subtracting recovered effects for each covariate from the variable of interest, thus changing the scale of the variable (y-axis). Significance of group differences (T-test) are indicated with star symbols, where ns: $2.38 \times 10^{-3} < p$, *: $2 \times 10^{-4} < p \le 2.38 \times 10^{-3}$, **: $2 \times 10^{-5} < p \le 2 \times 10^{-4}$, ***: $2 \times 10^{-6} < p \le 2 \times 10^{-5}$ ****: $p \le 2 \times 10^{-6}$.

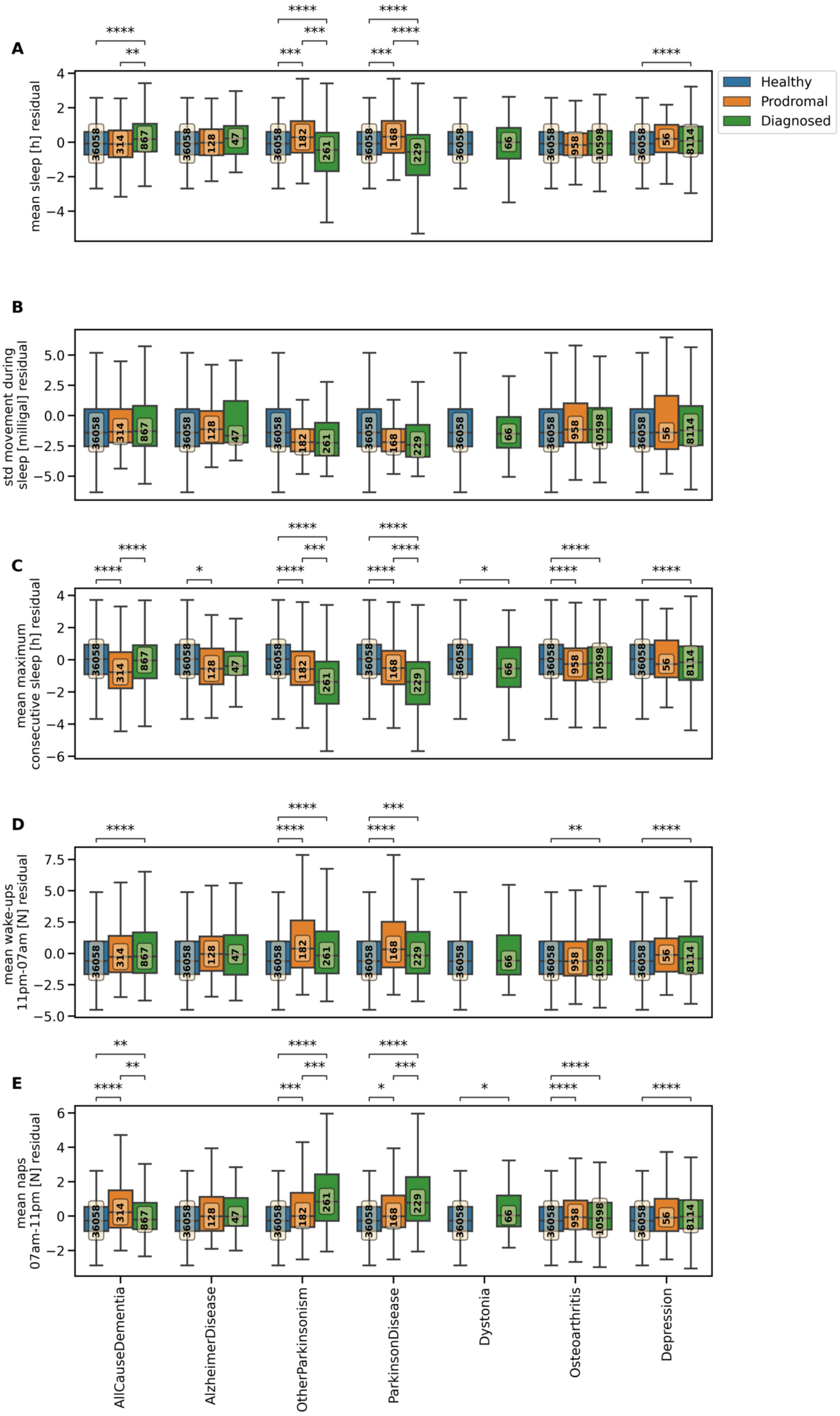

A
B
C
D
E
Healthy
Prodromal
Diagnosed
mean sleep [h] residual
std movement during sleep [milligal] residual
mean maximum consecutive sleep [h] residual
mean wake-ups 11pm-07am [N] residual
mean naps 07am-11pm [N] residual
AllCauseDementia
AlzheimerDisease
OtherParkinsonism
ParkinsonDisease
Dystonia
Osteoarthritis
Depression

##### Supplemental Figure 5: Quality and length of sleep in several disorders

Group comparisons between prodromal, diagnosed, and unaffected controls are shown before removal of comorbid depressed and PD subjects. The measures are age, BMI, and sex corrected through parameters learned from the unaffected population thus the residual is displayed and does not reflect the true value of the variable leading to potentially negative values. The boxplot shows the mean and quantiles. The yellow boxes show the number of subjects in each group. The asterisks indicate the significant difference between groups (<0.05 Bonferroni-corrected ns: $3.33x10^{-3} < p <= 1$, *: $3x10^{-4} < p <= 3.33x10^{-3}$, **: $3x10^{-5} < p <= 3x10^{-4}$, ***: $3x10^{-6} < p <= 3x10^{-5}$, ****: $p <= 3x10^{-6}$).

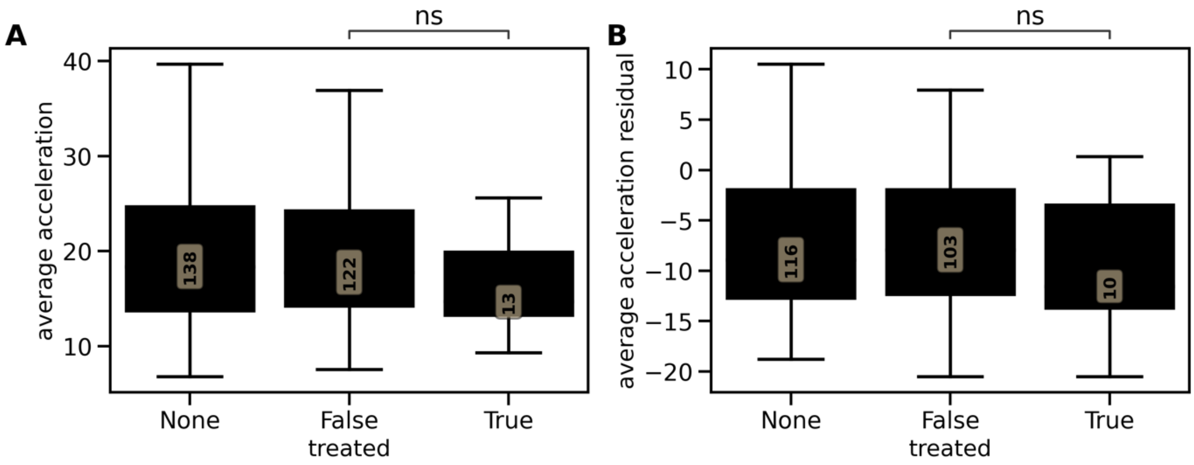


#### Supplemental Figure 6: Medication effect on acceleration

We plot the [A] average acceleration and the [B] residual average acceleration corrected for age, sex, and BMI as learned from unaffected controls for treated (N = 13, N = 10) vs non-treated diagnosed Parkinson's disease cases (N = 122, N = 103). Treated means here that no more than 10 weeks prior to accelerometry data collection a prescription for antiparkinsonism drugs was found in the associated primary care records. Subject with no GP prescription records available were excluded here (N = 138, N = 116).

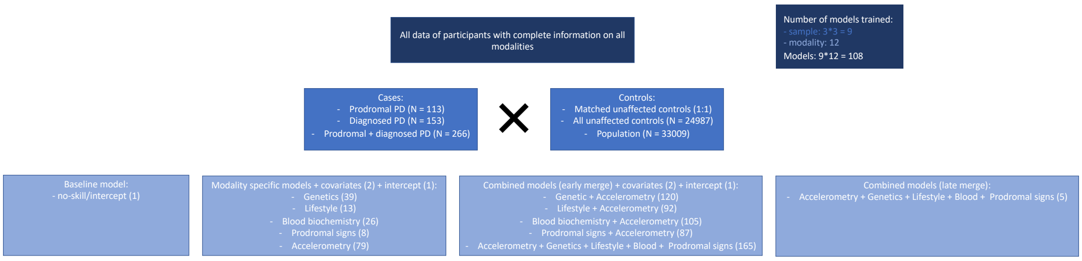


#### Supplemental Figure 7: Overview of all trained models

We show with which samples and which predictors each model is trained. With three different outcomes/cases and three different control groups, a total of nine different scenarios are modelled. We distinguish between baseline, modality-specific, and early and late combined models. For each model we show in brackets the number of predictors included.

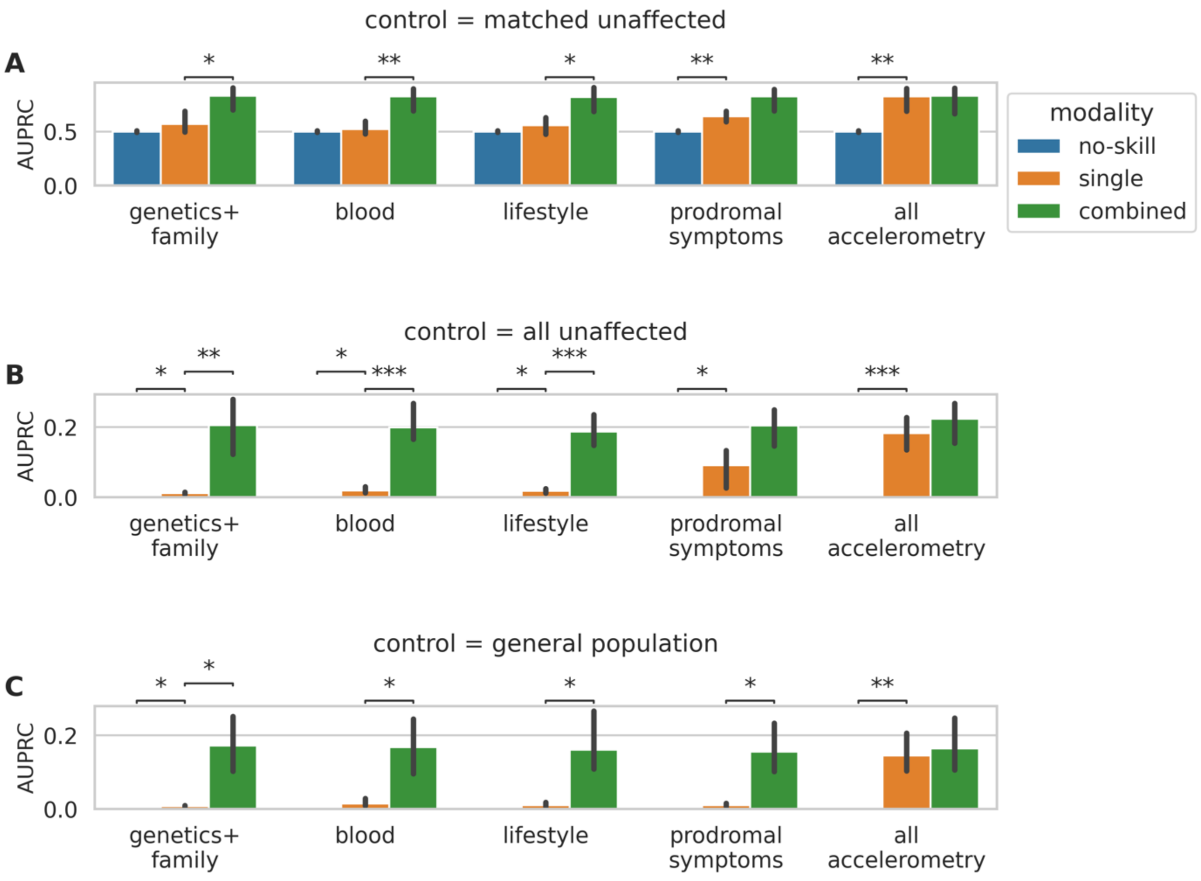


**Supplemental Figure 8: Model comparison to identify diagnosed Parkinson's disease**

The performance comparisons for each modality are shown. We compare no-skill (intercept) with single (modality-specific) and single with combined (modality + accelerometry modality). For the accelerometry modality, the combined model is the ones merging all modalities. We show this for each control group setting [A-C]. The barplot shows the mean performance across 5-folds and the error bars indicate the 95% Bonferroni-adjusted confidence interval. The asterisks indicate the significant difference between performances (<0.05 Bonferroni-corrected ns: $5x10^{-3} < p <=1$, *: $5x10^{-4} < p <=5x10^{-3}$, **: $5x10^{-5} < p <= 5x10^{-4}$, ***: $5x10^{-6} < p <= 5x10^{-5}$, ****: $p <= 5x10^{-6}$).

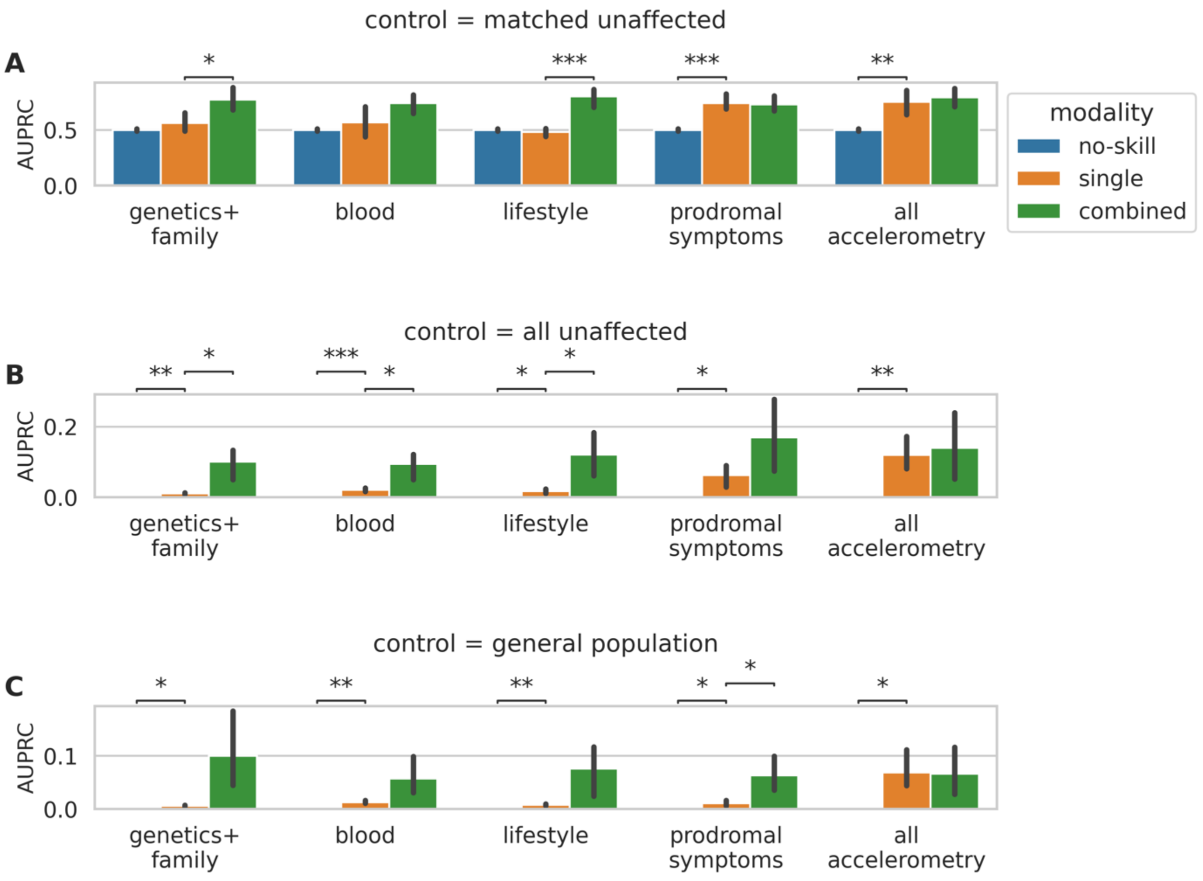


**Supplemental Figure 9: Model comparison to identify prodromal Parkinson's disease**

The performance comparisons for each modality are shown. We compare no-skill (intercept) with single (modality-specific) and single with combined (modality + accelerometry modality). For the accelerometry modality, the combined model is the ones merging all modalities. We show this for each control group setting [A-C]. The barplot shows the mean performance across 5-folds and the error bars indicate the 95% Bonferroni-adjusted confidence interval. The asterisks indicate the significant difference between performances (<0.05 Bonferroni-corrected ns: $5x10^{-3} < p <= 1$, *: $5x10^{-4} < p <= 5x10^{-3}$, **: $5x10^{-5} < p <= 5x10^{-4}$, ***: $5x10^{-6} < p <= 5x10^{-5}$, ****: $p <= 5x10^{-6}$).

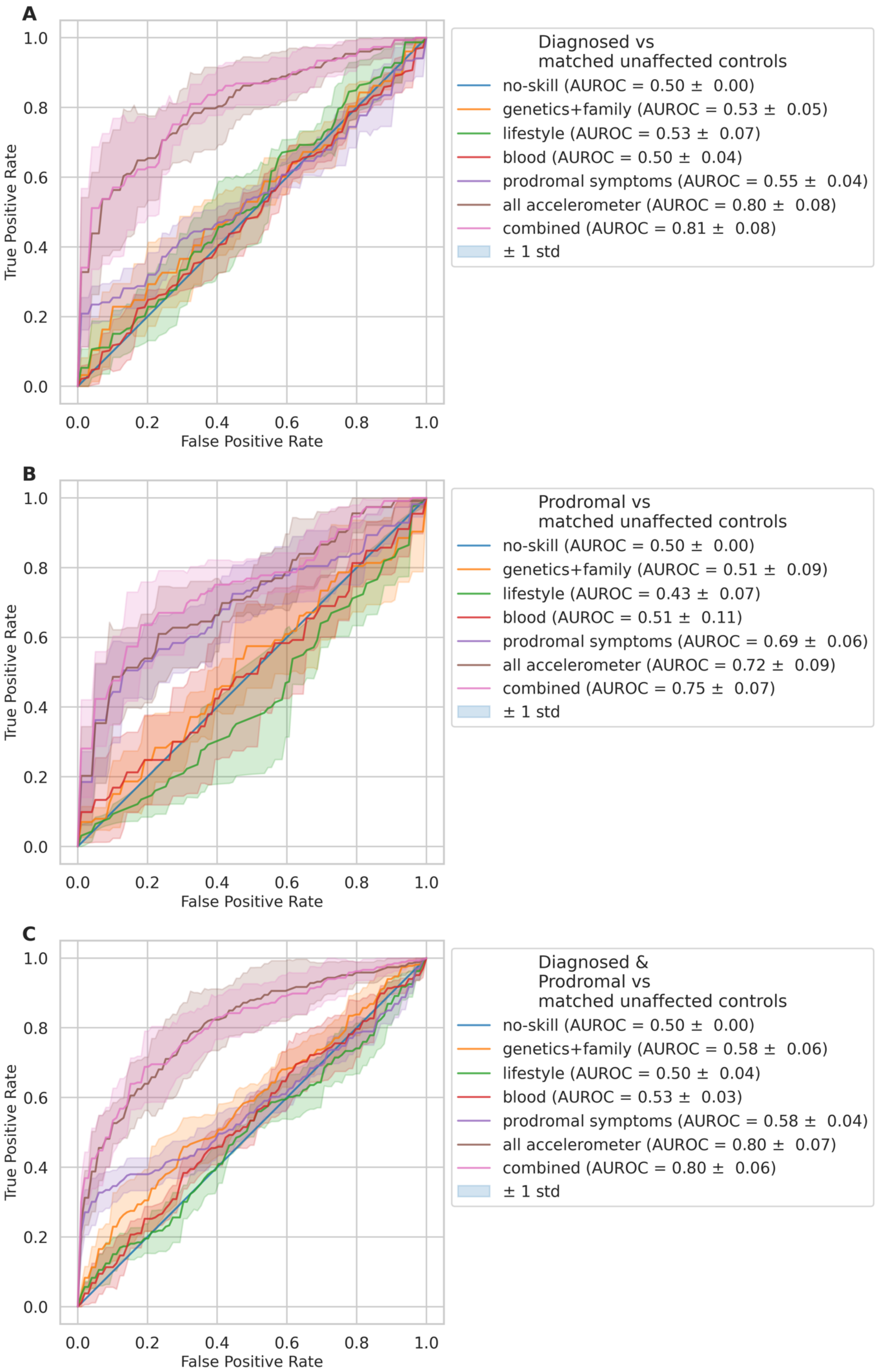

A
1.0
0.8
0.6
0.4
0.2
0.0
True Positive Rate
0.0
0.2
0.4
0.6
0.8
1.0
False Positive Rate
Diagnosed vs
matched unaffected controls
no-skill (AUROC = 0.50 ± 0.00)
genetics+family (AUROC = 0.53 ± 0.05)
lifestyle (AUROC = 0.53 ± 0.07)
blood (AUROC = 0.50 ± 0.04)
prodromal symptoms (AUROC = 0.55 ± 0.04)
all accelerometer (AUROC = 0.80 ± 0.08)
combined (AUROC = 0.81 ± 0.08)
± 1 std
B
1.0
0.8
0.6
0.4
0.2
0.0
True Positive Rate
0.0
0.2
0.4
0.6
0.8
1.0
False Positive Rate
Prodromal vs
matched unaffected controls
no-skill (AUROC = 0.50 ± 0.00)
genetics+family (AUROC = 0.51 ± 0.09)
lifestyle (AUROC = 0.43 ± 0.07)
blood (AUROC = 0.51 ± 0.11)
prodromal symptoms (AUROC = 0.69 ± 0.06)
all accelerometer (AUROC = 0.72 ± 0.09)
combined (AUROC = 0.75 ± 0.07)
± 1 std
C
1.0
0.8
0.6
0.4
0.2
0.0
True Positive Rate
0.0
0.2
0.4
0.6
0.8
1.0
False Positive Rate
Diagnosed &
Prodromal vs
matched unaffected controls
no-skill (AUROC = 0.50 ± 0.00)
genetics+family (AUROC = 0.58 ± 0.06)
lifestyle (AUROC = 0.50 ± 0.04)
blood (AUROC = 0.53 ± 0.03)
prodromal symptoms (AUROC = 0.58 ± 0.04)
all accelerometer (AUROC = 0.80 ± 0.07)
combined (AUROC = 0.80 ± 0.06)
± 1 std

#### Supplemental Figure 10: Receiver operator curves for matched healthy control models

The mean receiver operator curves across the outer five-folds are shown together with their standard deviation. We show this for each model type: [A] identifying diseased cases, [B] identifying prodromal cases, [C] identifying diseased and prodromal cases from matched healthy controls for each tested modality.

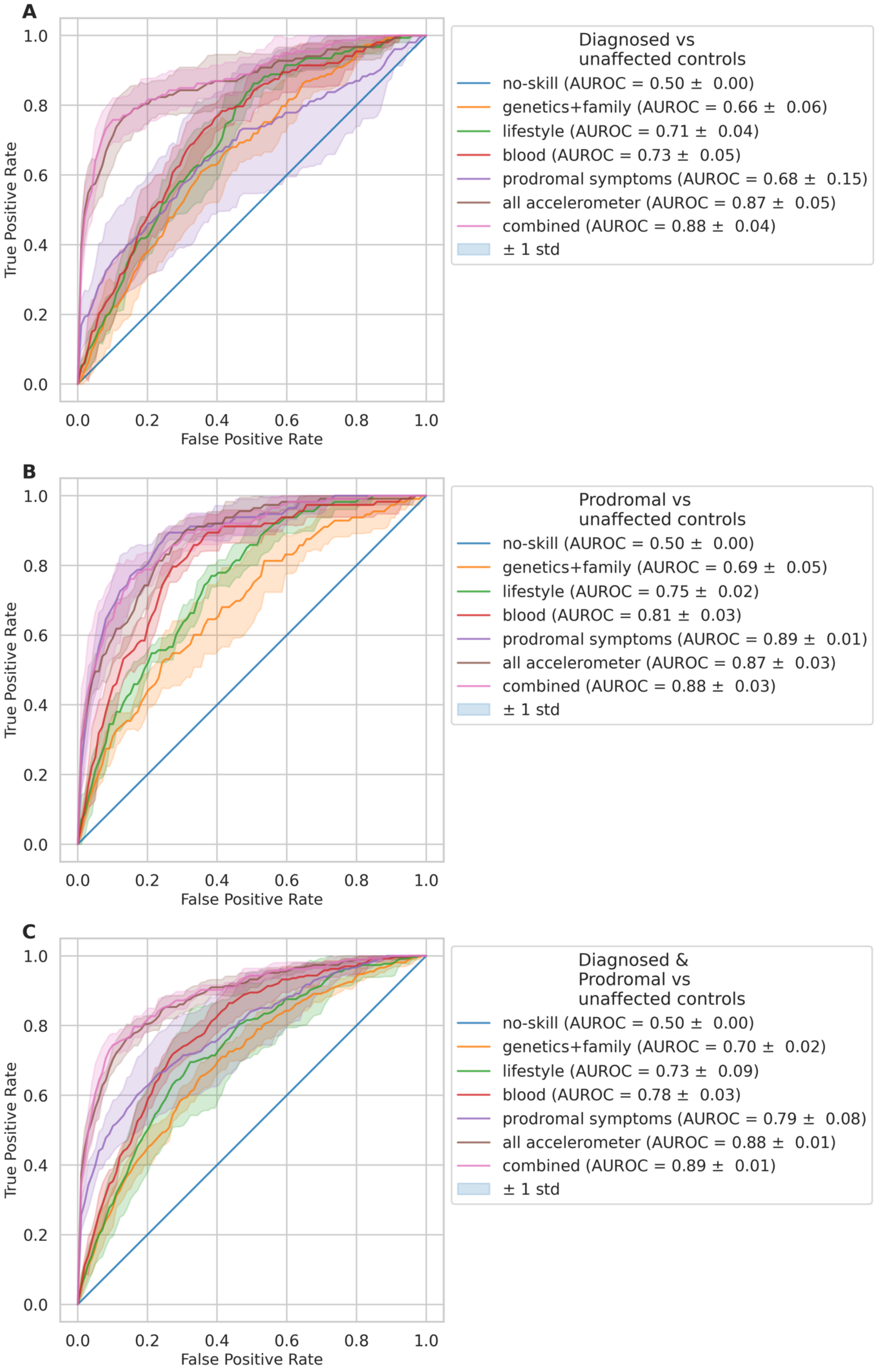
A
Diagnosed vs unaffected controls
no-skill (AUROC = 0.50 ± 0.00)
genetics+family (AUROC = 0.66 ± 0.06)
lifestyle (AUROC = 0.71 ± 0.04)
blood (AUROC = 0.73 ± 0.05)
prodromal symptoms (AUROC = 0.68 ± 0.15)
all accelerometer (AUROC = 0.87 ± 0.05)
combined (AUROC = 0.88 ± 0.04)
± 1 std
True Positive Rate
False Positive Rate
B
Prodromal vs unaffected controls
no-skill (AUROC = 0.50 ± 0.00)
genetics+family (AUROC = 0.69 ± 0.05)
lifestyle (AUROC = 0.75 ± 0.02)
blood (AUROC = 0.81 ± 0.03)
prodromal symptoms (AUROC = 0.89 ± 0.01)
all accelerometer (AUROC = 0.87 ± 0.03)
combined (AUROC = 0.88 ± 0.03)
± 1 std
True Positive Rate
False Positive Rate
C
Diagnosed & Prodromal vs unaffected controls
no-skill (AUROC = 0.50 ± 0.00)
genetics+family (AUROC = 0.70 ± 0.02)
lifestyle (AUROC = 0.73 ± 0.09)
blood (AUROC = 0.78 ± 0.03)
prodromal symptoms (AUROC = 0.79 ± 0.08)
all accelerometer (AUROC = 0.88 ± 0.01)
combined (AUROC = 0.89 ± 0.01)
± 1 std
True Positive Rate
False Positive Rate

#### Supplemental Figure 11: Receiver operator curves for unmatched healthy control models

The mean receiver operator curves across the outer five-folds are shown together with their standard deviation. We show this for each model type: [A] identifying diseased cases, [B] identifying prodromal cases, [C] identifying diseased and prodromal cases from unmatched healthy controls for each tested modality.

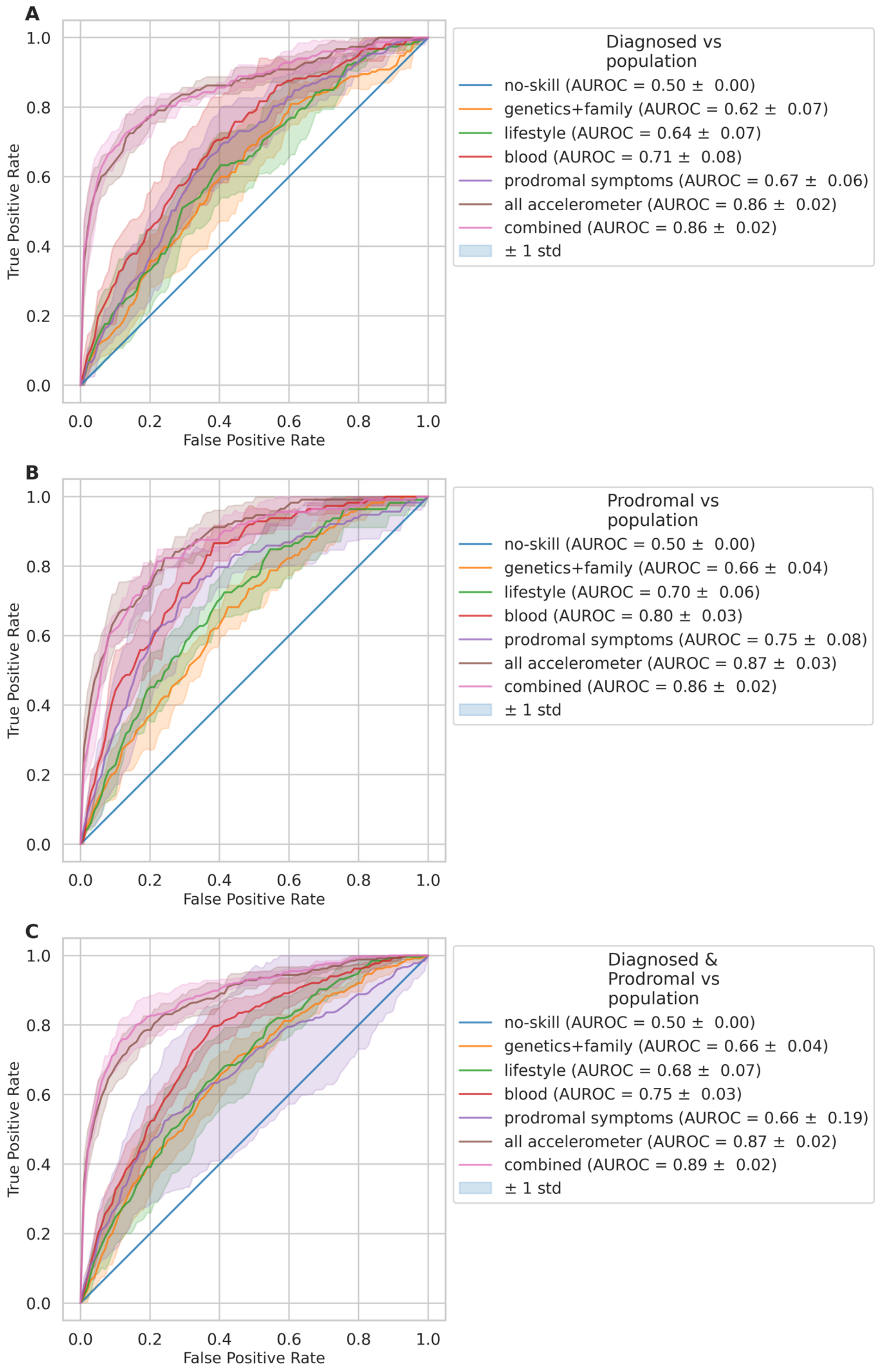
A
Diagnosed vs population
no-skill (AUROC = 0.50 ± 0.00)
genetics+family (AUROC = 0.62 ± 0.07)
lifestyle (AUROC = 0.64 ± 0.07)
blood (AUROC = 0.71 ± 0.08)
prodromal symptoms (AUROC = 0.67 ± 0.06)
all accelerometer (AUROC = 0.86 ± 0.02)
combined (AUROC = 0.86 ± 0.02)
± 1 std
True Positive Rate
False Positive Rate
0.0
0.2
0.4
0.6
0.8
1.0
B
Prodromal vs population
no-skill (AUROC = 0.50 ± 0.00)
genetics+family (AUROC = 0.66 ± 0.04)
lifestyle (AUROC = 0.70 ± 0.06)
blood (AUROC = 0.80 ± 0.03)
prodromal symptoms (AUROC = 0.75 ± 0.08)
all accelerometer (AUROC = 0.87 ± 0.03)
combined (AUROC = 0.86 ± 0.02)
± 1 std
True Positive Rate
False Positive Rate
C
Diagnosed & Prodromal vs population
no-skill (AUROC = 0.50 ± 0.00)
genetics+family (AUROC = 0.66 ± 0.04)
lifestyle (AUROC = 0.68 ± 0.07)
blood (AUROC = 0.75 ± 0.03)
prodromal symptoms (AUROC = 0.66 ± 0.19)
all accelerometer (AUROC = 0.87 ± 0.02)
combined (AUROC = 0.89 ± 0.02)
± 1 std
True Positive Rate
False Positive Rate

#### Supplemental Figure 12: Receiver operator curves for population-based models

The mean receiver operator curves across the outer five-folds are shown together with their standard deviation. We show this for each model type: [A] identifying diseased cases, [B] identifying prodromal cases, [C] identifying diseased and prodromal cases from the population for each tested modality.

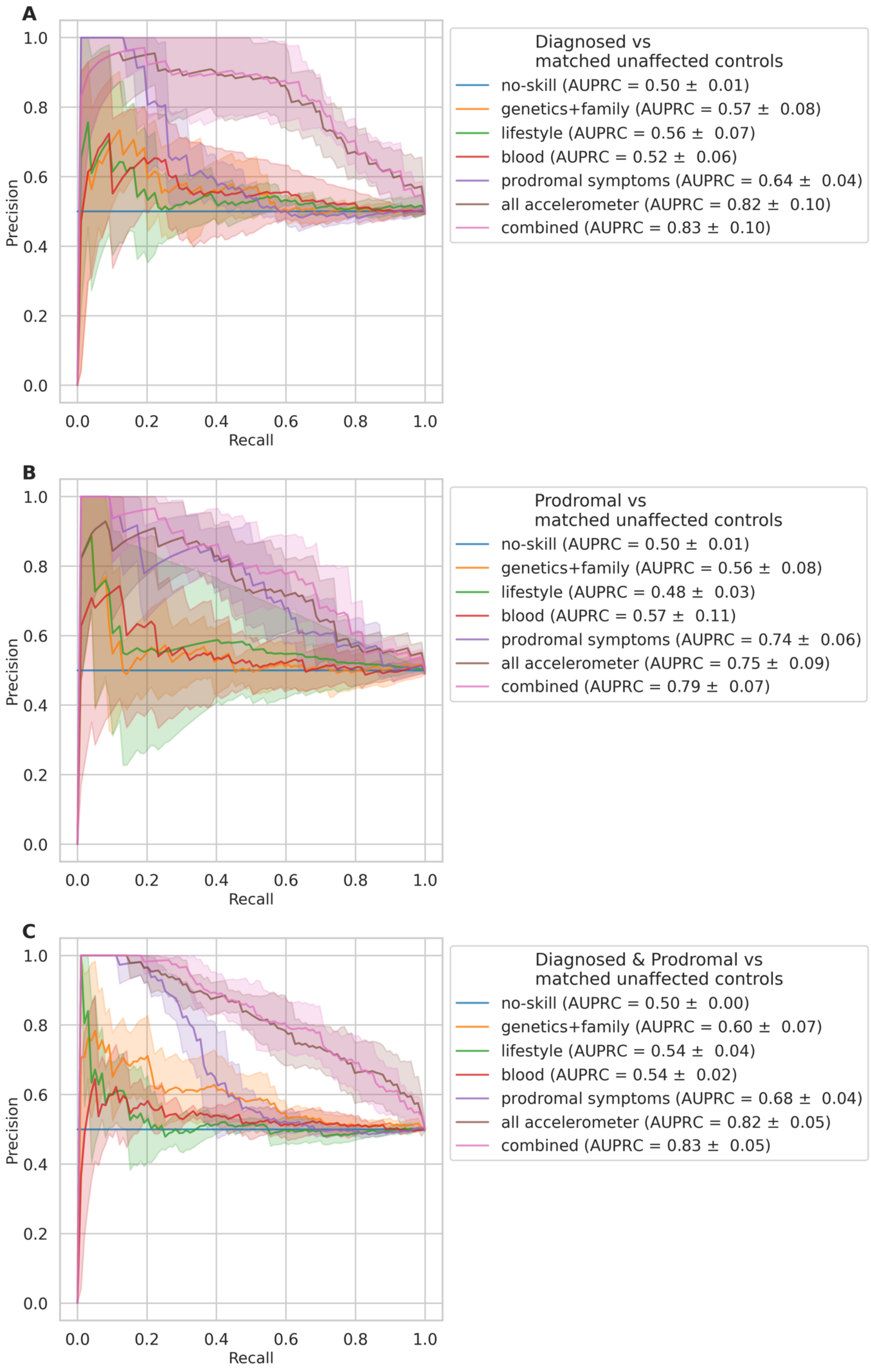
A
Diagnosed vs
matched unaffected controls
no-skill (AUPRC = 0.50 ± 0.01)
genetics+family (AUPRC = 0.57 ± 0.08)
lifestyle (AUPRC = 0.56 ± 0.07)
blood (AUPRC = 0.52 ± 0.06)
prodromal symptoms (AUPRC = 0.64 ± 0.04)
all accelerometer (AUPRC = 0.82 ± 0.10)
combined (AUPRC = 0.83 ± 0.10)
Precision
Recall
0.0
0.2
0.4
0.6
0.8
1.0
B
Prodromal vs
matched unaffected controls
no-skill (AUPRC = 0.50 ± 0.01)
genetics+family (AUPRC = 0.56 ± 0.08)
lifestyle (AUPRC = 0.48 ± 0.03)
blood (AUPRC = 0.57 ± 0.11)
prodromal symptoms (AUPRC = 0.74 ± 0.06)
all accelerometer (AUPRC = 0.75 ± 0.09)
combined (AUPRC = 0.79 ± 0.07)
Precision
Recall
C
Diagnosed & Prodromal vs
matched unaffected controls
no-skill (AUPRC = 0.50 ± 0.00)
genetics+family (AUPRC = 0.60 ± 0.07)
lifestyle (AUPRC = 0.54 ± 0.04)
blood (AUPRC = 0.54 ± 0.02)
prodromal symptoms (AUPRC = 0.68 ± 0.04)
all accelerometer (AUPRC = 0.82 ± 0.05)
combined (AUPRC = 0.83 ± 0.05)
Precision
Recall

#### Supplemental Figure 13: Precision recall curves for matched healthy control models

The mean precision recall curves across the outer five-folds are shown together with their standard deviation. We show this for each model type: [A] identifying diseased cases, [B] identifying prodromal cases, [C] identifying diseased and prodromal cases from matched healthy controls for each tested modality.

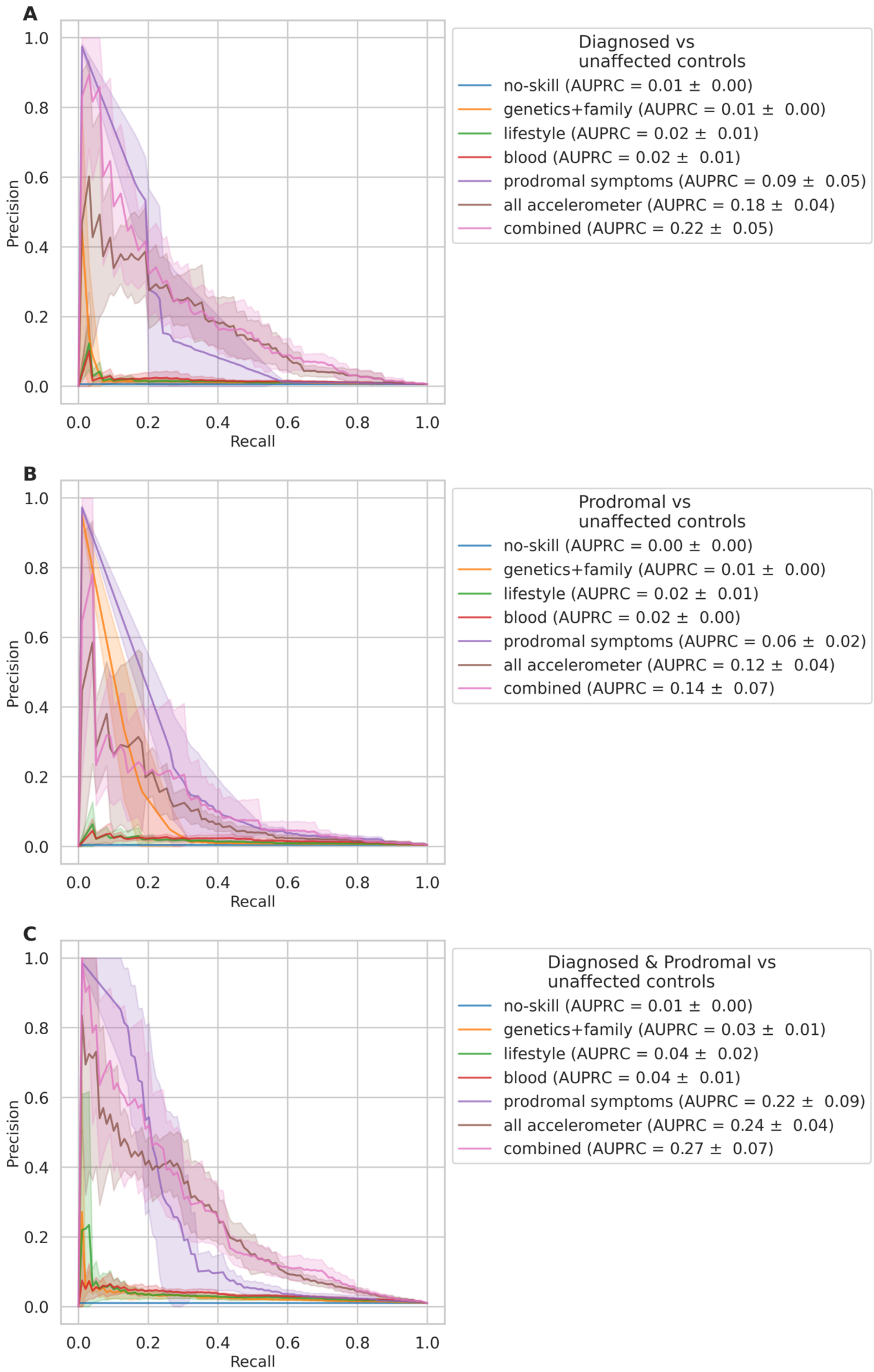
A
Diagnosed vs unaffected controls
no-skill (AUPRC = 0.01 ± 0.00)
genetics+family (AUPRC = 0.01 ± 0.00)
lifestyle (AUPRC = 0.02 ± 0.01)
blood (AUPRC = 0.02 ± 0.01)
prodromal symptoms (AUPRC = 0.09 ± 0.05)
all accelerometer (AUPRC = 0.18 ± 0.04)
combined (AUPRC = 0.22 ± 0.05)
Precision
Recall
B
Prodromal vs unaffected controls
no-skill (AUPRC = 0.00 ± 0.00)
genetics+family (AUPRC = 0.01 ± 0.00)
lifestyle (AUPRC = 0.02 ± 0.01)
blood (AUPRC = 0.02 ± 0.00)
prodromal symptoms (AUPRC = 0.06 ± 0.02)
all accelerometer (AUPRC = 0.12 ± 0.04)
combined (AUPRC = 0.14 ± 0.07)
Precision
Recall
C
Diagnosed & Prodromal vs unaffected controls
no-skill (AUPRC = 0.01 ± 0.00)
genetics+family (AUPRC = 0.03 ± 0.01)
lifestyle (AUPRC = 0.04 ± 0.02)
blood (AUPRC = 0.04 ± 0.01)
prodromal symptoms (AUPRC = 0.22 ± 0.09)
all accelerometer (AUPRC = 0.24 ± 0.04)
combined (AUPRC = 0.27 ± 0.07)
Precision
Recall

#### Supplemental Figure 14: Precision recall curves for unmatched healthy control models

The mean precision recall curves across the outer five-folds are shown together with their standard deviation. We show this for each model type: [A] identifying diseased cases, [B] identifying prodromal cases, [C] identifying diseased and prodromal cases from unmatched healthy controls for each tested modality.

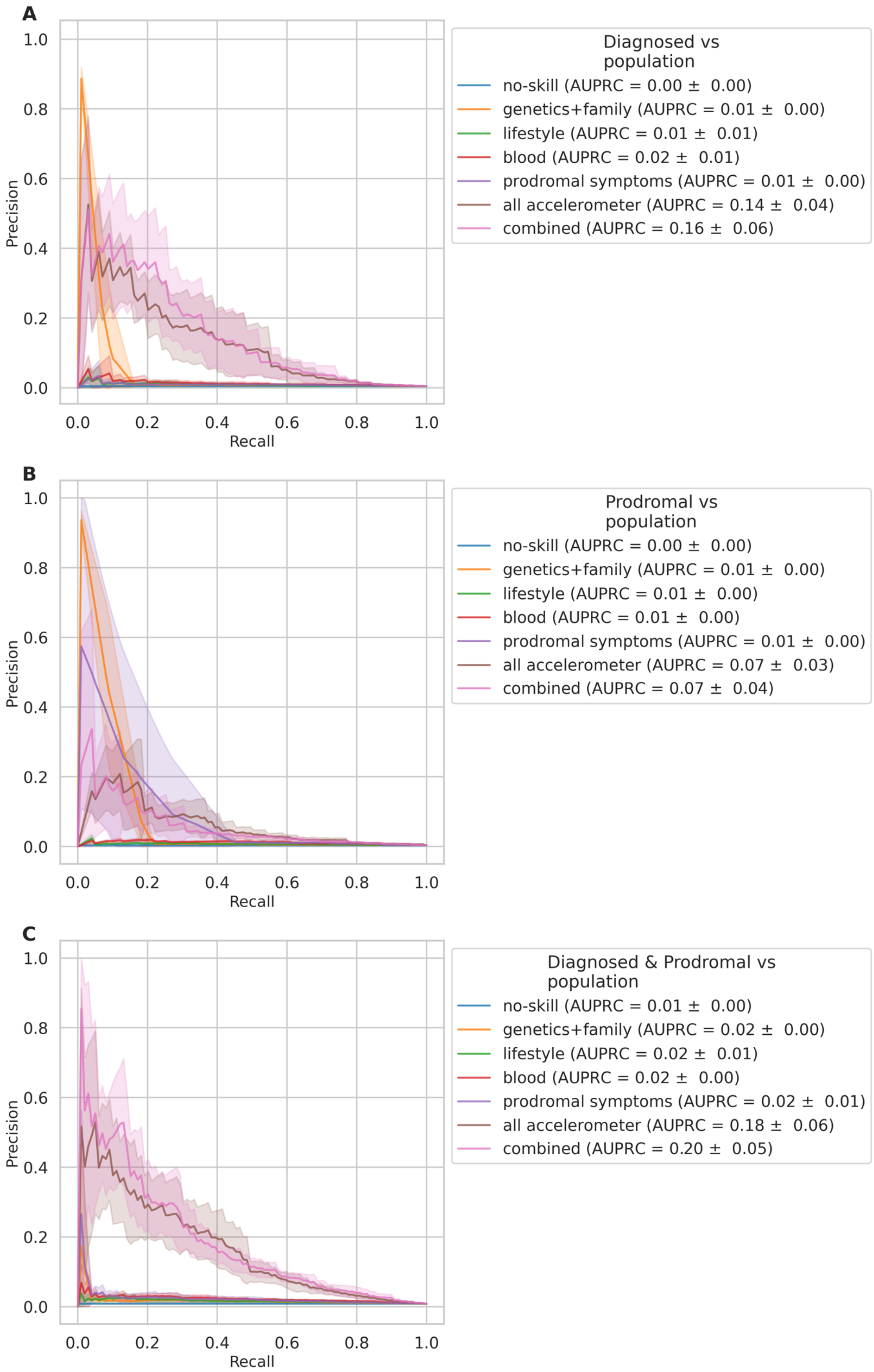
A
Diagnosed vs population
no-skill (AUPRC = 0.00 ± 0.00)
genetics+family (AUPRC = 0.01 ± 0.00)
lifestyle (AUPRC = 0.01 ± 0.01)
blood (AUPRC = 0.02 ± 0.01)
prodromal symptoms (AUPRC = 0.01 ± 0.00)
all accelerometer (AUPRC = 0.14 ± 0.04)
combined (AUPRC = 0.16 ± 0.06)
Precision
Recall
B
Prodromal vs population
no-skill (AUPRC = 0.00 ± 0.00)
genetics+family (AUPRC = 0.01 ± 0.00)
lifestyle (AUPRC = 0.01 ± 0.00)
blood (AUPRC = 0.01 ± 0.00)
prodromal symptoms (AUPRC = 0.01 ± 0.00)
all accelerometer (AUPRC = 0.07 ± 0.03)
combined (AUPRC = 0.07 ± 0.04)
Precision
Recall
C
Diagnosed & Prodromal vs population
no-skill (AUPRC = 0.01 ± 0.00)
genetics+family (AUPRC = 0.02 ± 0.00)
lifestyle (AUPRC = 0.02 ± 0.01)
blood (AUPRC = 0.02 ± 0.00)
prodromal symptoms (AUPRC = 0.02 ± 0.01)
all accelerometer (AUPRC = 0.18 ± 0.06)
combined (AUPRC = 0.20 ± 0.05)
Precision
Recall

#### Supplemental Figure 15: Precision recall curves for population-based models

The mean precision recall curves across the outer five-folds are shown together with their standard deviation. We show this for each model type: [A] identifying diseased cases, [B] identifying prodromal cases, [C] identifying diseased and prodromal cases from the population for each tested modality.

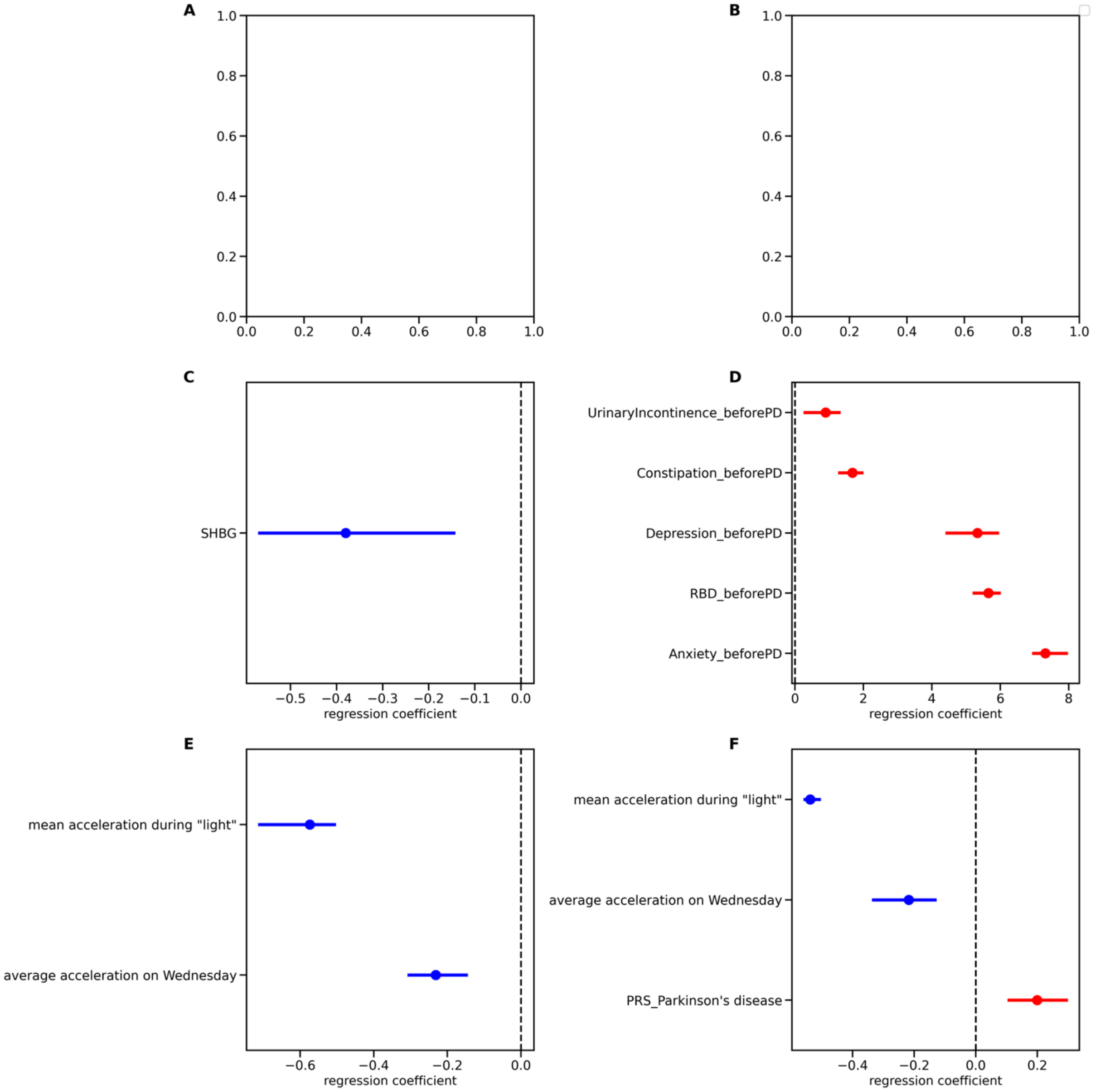


**Supplemental Figure 16: Feature importance for each modality for the prodromal vs matched unaffected controls models**

The most important, significant features across the cross-validation splits are shown for the prodromal Parkinson's disease (PD) vs matched unaffected controls models. For each feature we show the regression coefficient and the 95% CI corrected for multiple comparisons with the Bonferroni-correction. Features that increase the likelihood of getting PD are shown in red, whereas those that decrease the likelihood are shown in blue. Each plot shows the features of a model trained on a different feature set/modality: [A] genetics+family, [B] lifestyle, [C] blood, [D] prodromal symptoms, [E] all accelerometry, [F] combined.

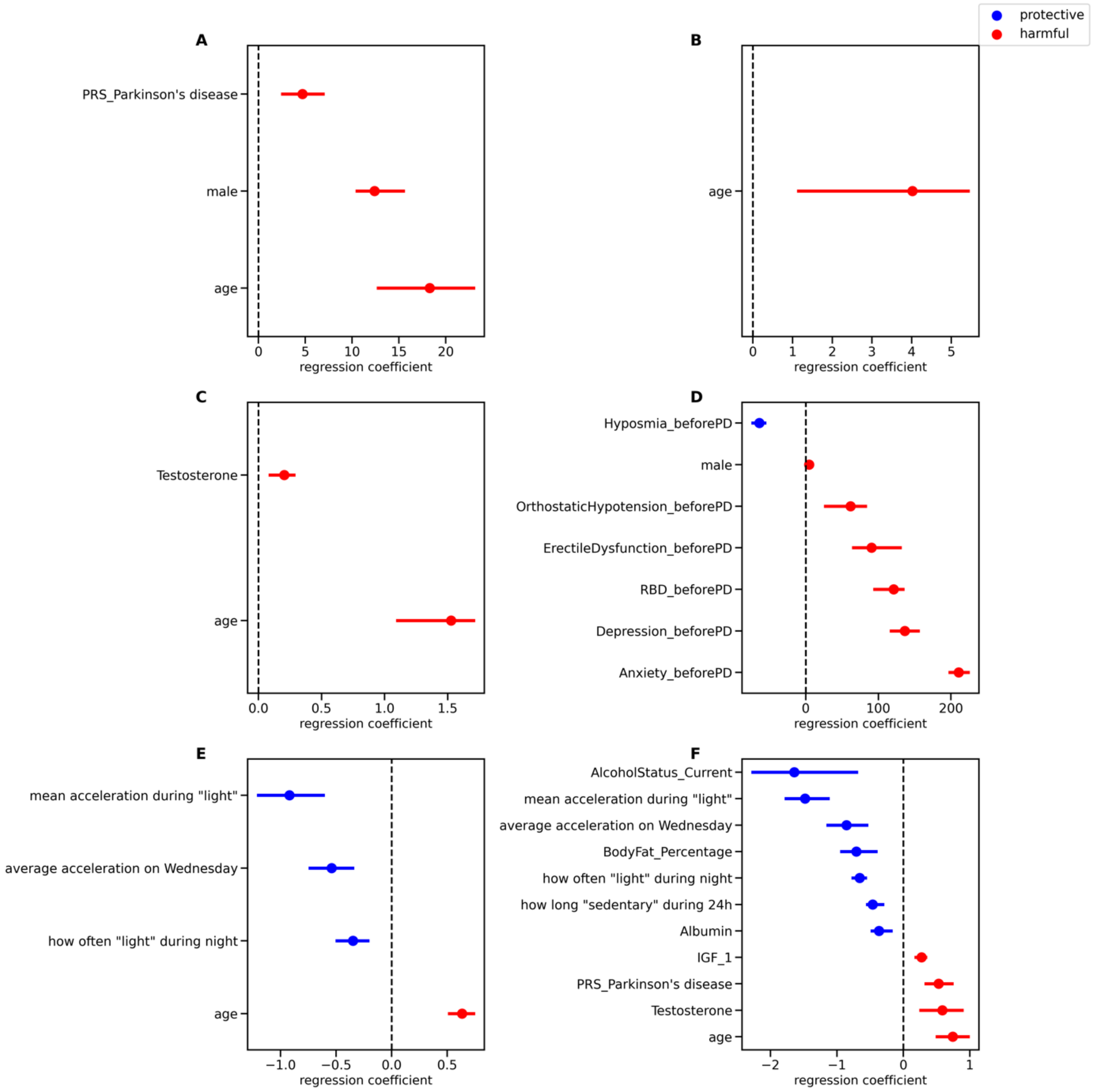


#### Supplemental Figure 17: Feature importance for each modality for the prodromal vs all unaffected controls models

The most important, significant features across the cross-validation splits are shown for the prodromal Parkinson's disease (PD) vs matched unaffected controls models. For each feature we show the regression coefficient and the 95% CI corrected for multiple comparisons with the Bonferroni-correction. Features that increase the likelihood of getting PD are shown in red, whereas those that decrease the likelihood are shown in blue. Each plot shows the features of a model trained on a different feature set/modality: [A] genetics+family, [B] lifestyle, [C] blood, [D] prodromal symptoms, [E] all accelerometry, [F] combined.

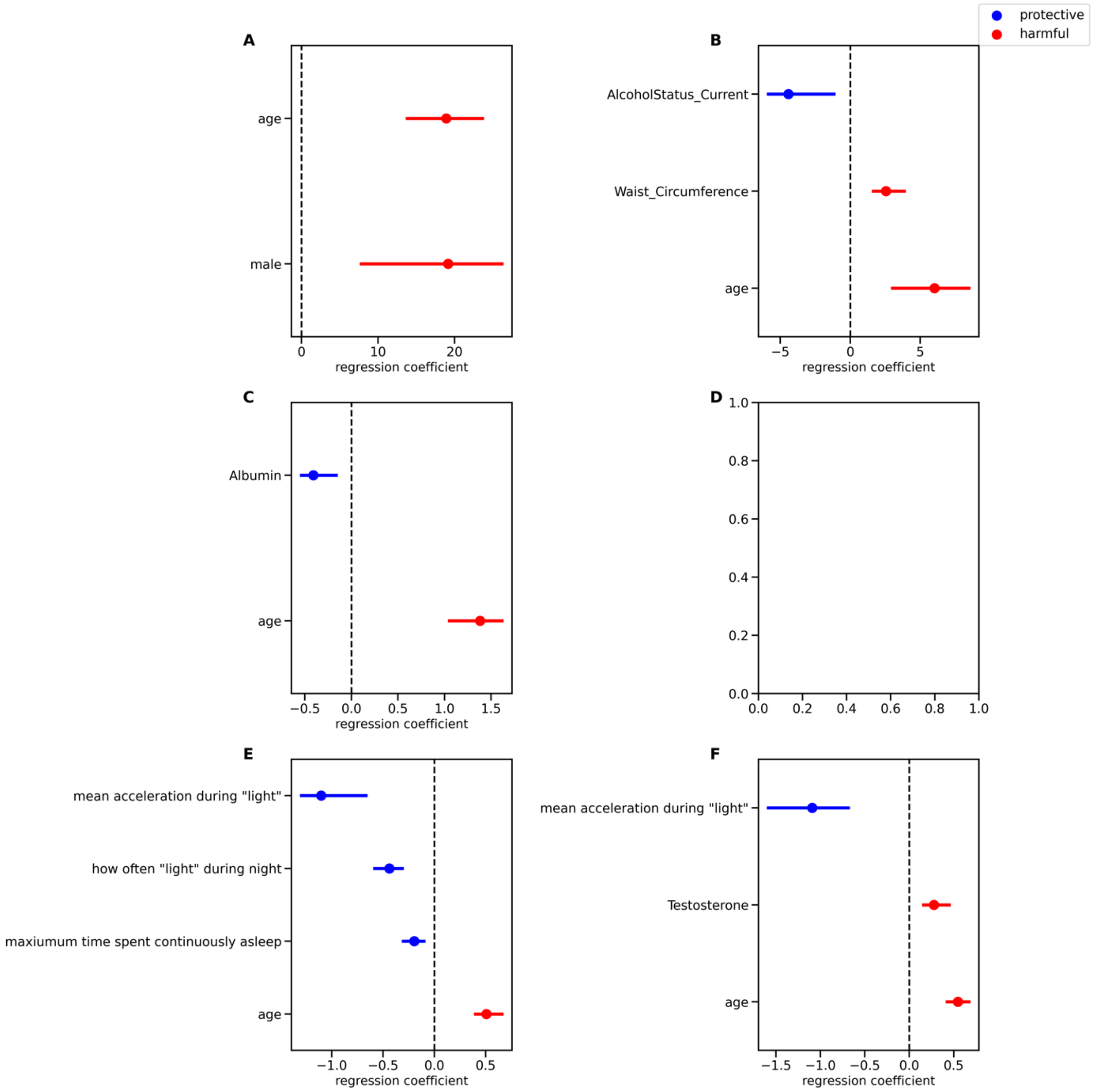


**Supplemental Figure 18: Feature importance for each modality for the prodromal vs population models**

The most important, significant features across the cross-validation splits are shown for the prodromal Parkinson's disease (PD) vs matched unaffected controls models. For each feature we show the regression coefficient and the 95% CI corrected for multiple comparisons with the Bonferroni-correction. Features that increase the likelihood of getting PD are shown in red, whereas those that decrease the likelihood are shown in blue. Each plot shows the features of a model trained on a different feature set/modality: [A] genetics+family, [B] lifestyle, [C] blood, [D] prodromal symptoms, [E] all accelerometry, [F] combined.

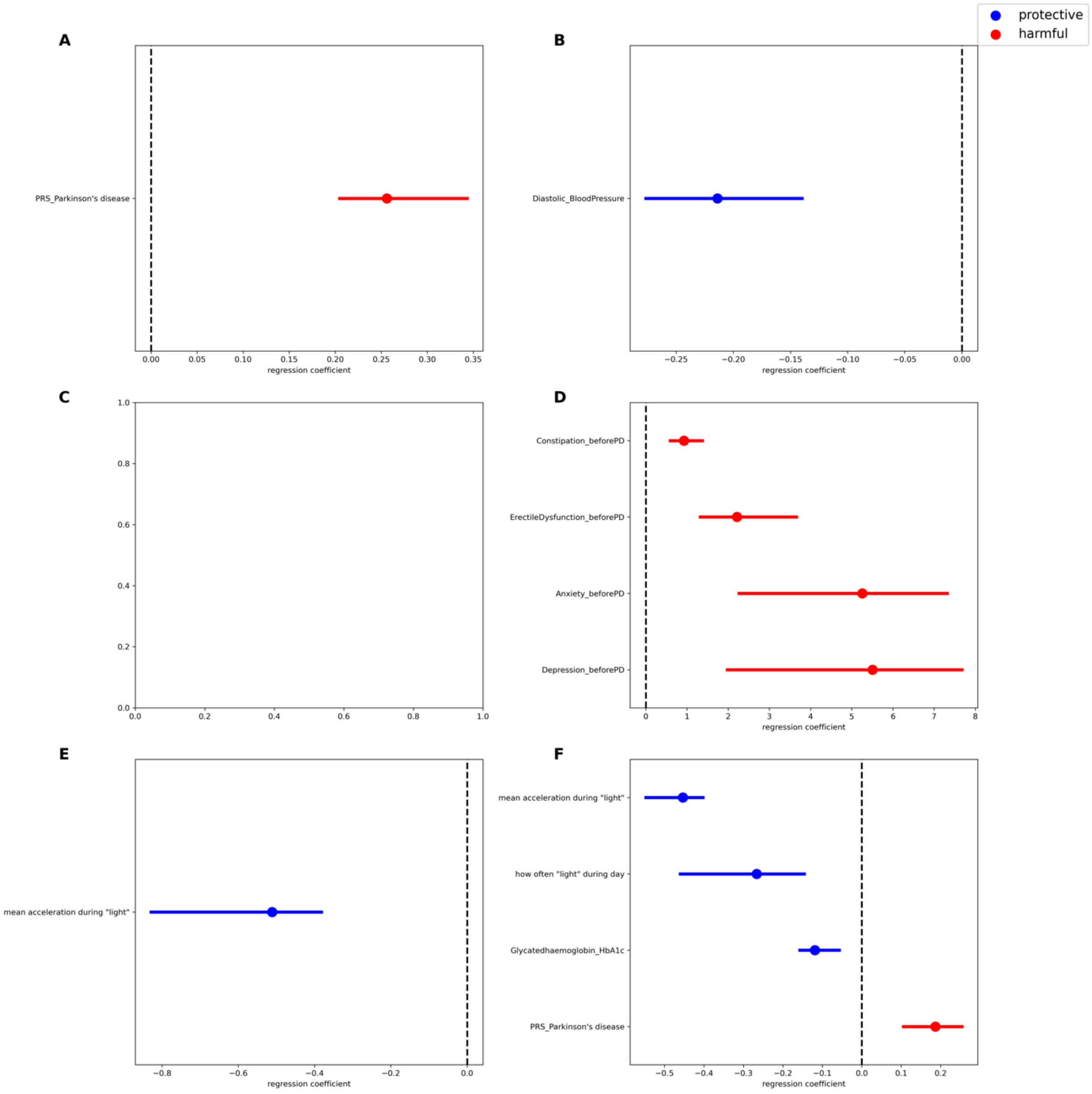


#### Supplemental Figure 19: Feature importance for each modality for the diagnosed vs matched unaffected controls models

The most important, significant features across the cross-validation splits are shown for the diagnosed Parkinson's disease (PD) vs matched unaffected controls models. For each feature we show the regression coefficient and the 95% CI corrected for multiple comparisons with the Bonferroni-correction. Features that increase the likelihood of having PD are shown in red, whereas those that decrease the likelihood are shown in blue. Each plot shows the features of a model trained on a different feature set/modality: [A] genetics+family, [B] lifestyle, [C] blood, [D] prodromal symptoms, [E] all accelerometry, [F] combined.

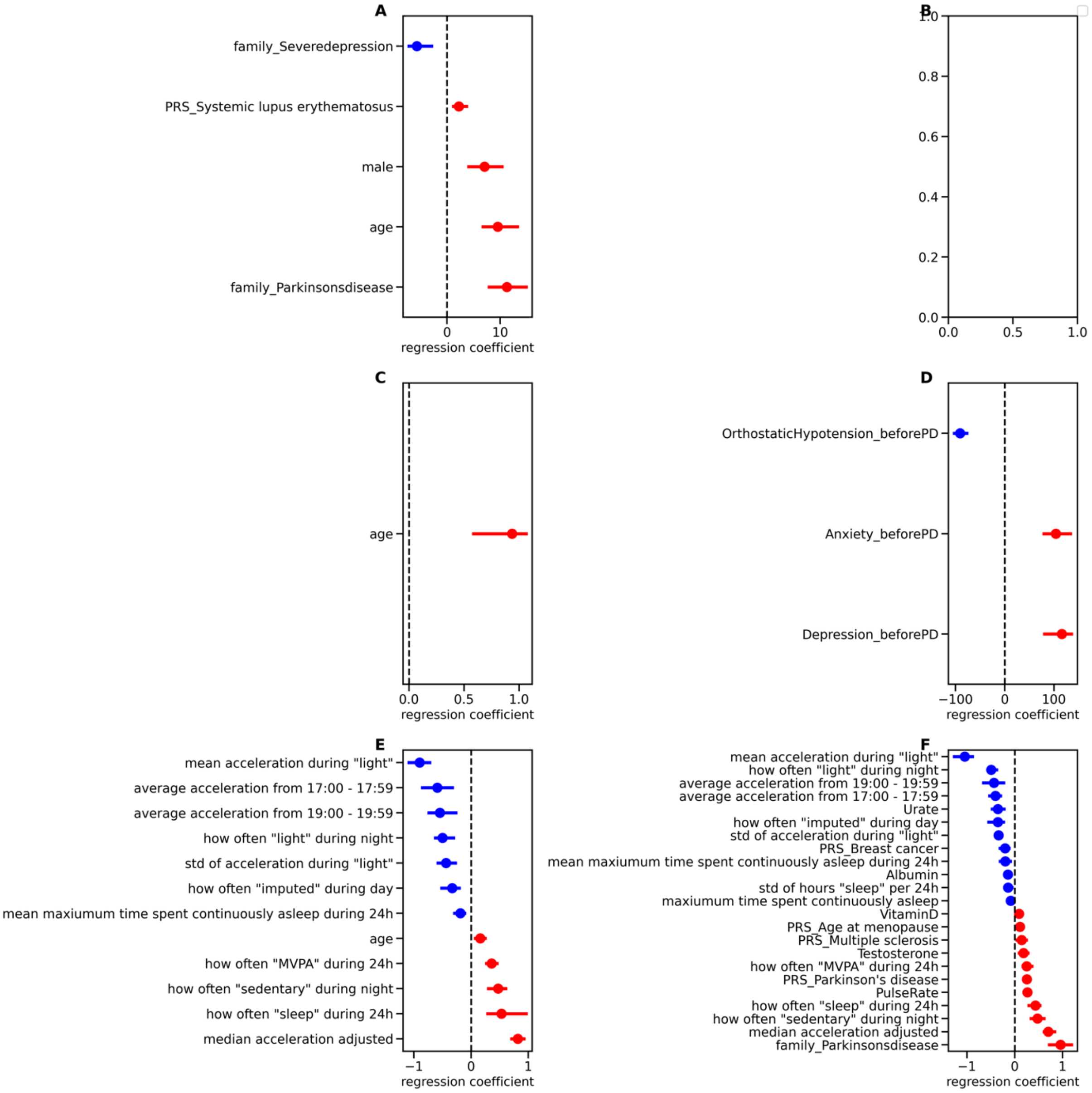


#### Supplemental Figure 20: Feature importance for each modality for the diagnosed vs all unaffected controls models

The most important, significant features across the cross-validation splits are shown for the diagnosed Parkinson's disease (PD) vs matched unaffected controls models. For each feature we show the regression coefficient and the 95% CI corrected for multiple comparisons with the Bonferroni-correction. Features that increase the likelihood of having PD are shown in red, whereas those that decrease the likelihood are shown in blue. Each subplot shows the features of one model trained on a different feature set/modality: [A] genetics+family, [B] lifestyle, [C] blood, [D] prodromal symptoms, [E] all accelerometry, [F] combined.

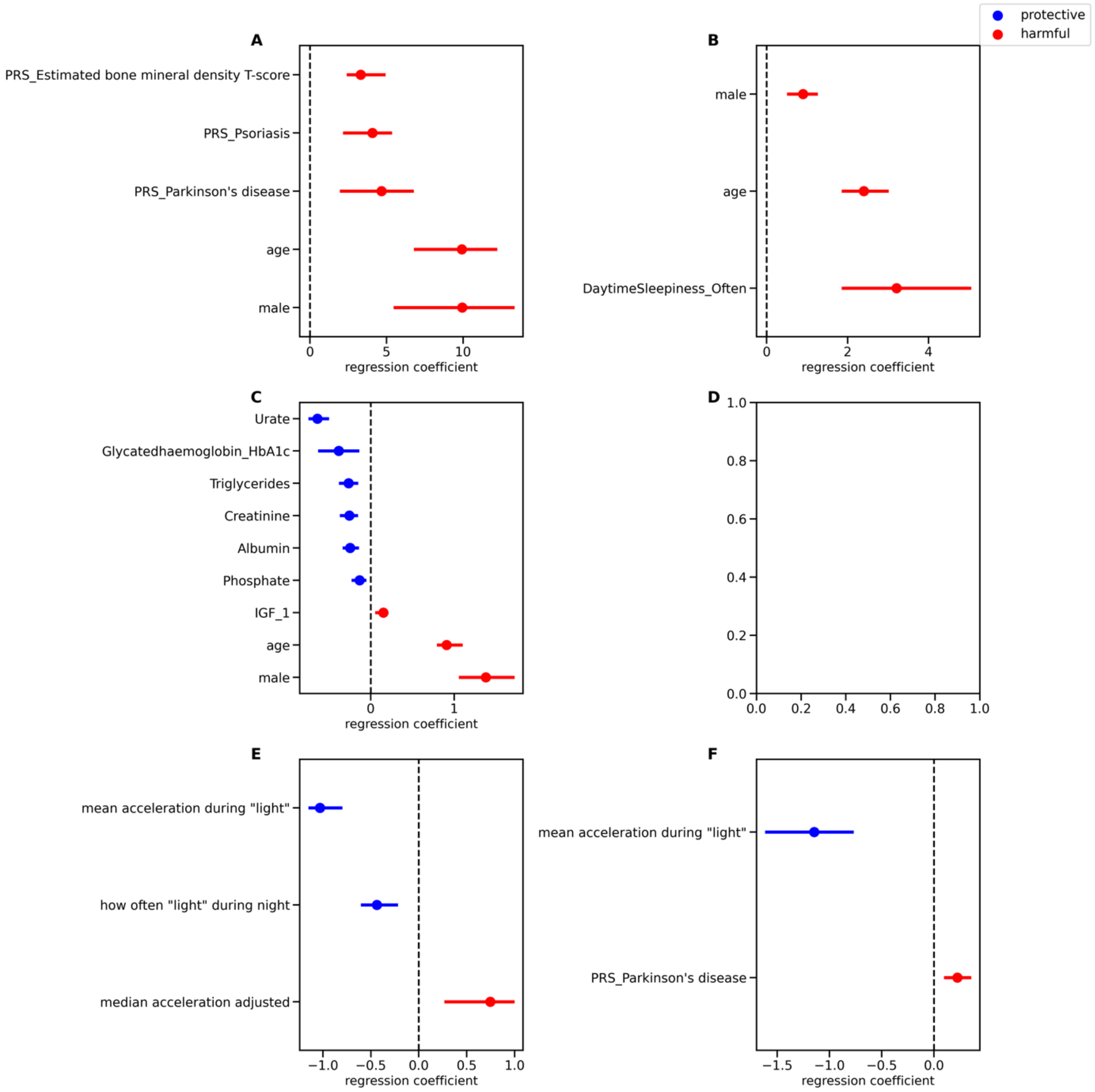


**Supplemental Figure 21: Feature importance for each modality for the diagnosed vs population models**

The most important, significant features across the cross-validation splits are shown for the diagnosed Parkinson's disease (PD) vs matched unaffected controls models. For each feature we show the regression coefficient and the 95% CI corrected for multiple comparisons with the Bonferroni-correction. Features that increase the likelihood of having PD are shown in red, whereas those that decrease the likelihood are shown in blue. Each plot shows the features of a model trained on a different feature set/modality: [A] genetics+family, [B] lifestyle, [C] blood, [D] prodromal symptoms, [E] all accelerometry, [F] combined.

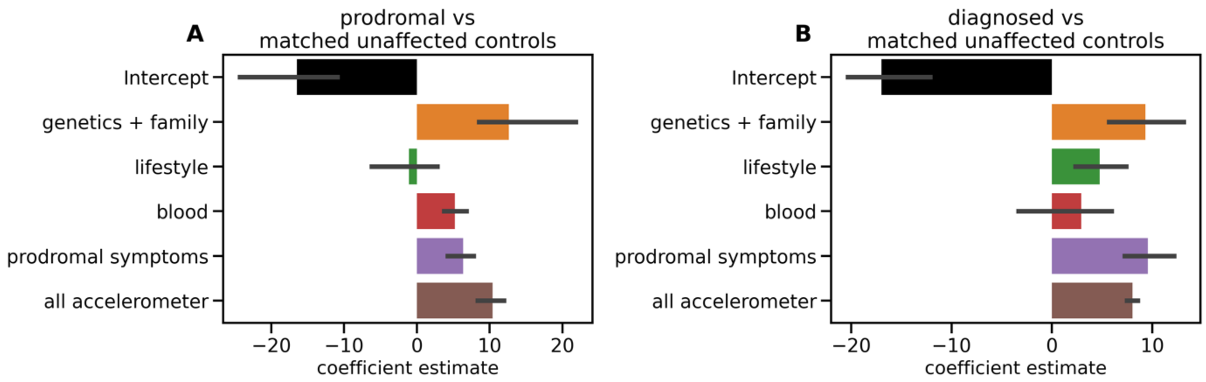


#### Supplemental Figure 22: Modality importance for the stacked model

The coefficients assigned to the modality specific predictions are shown. We plot the mean coefficient across folds and the Bonferroni-adjusted 95% Confidence Interval across those five folds. [A] shows the coefficients of the prodromal Parkinson's disease (PD) vs matched unaffected controls model and [B] the ones of the diagnosed PD vs matched unaffected controls model.

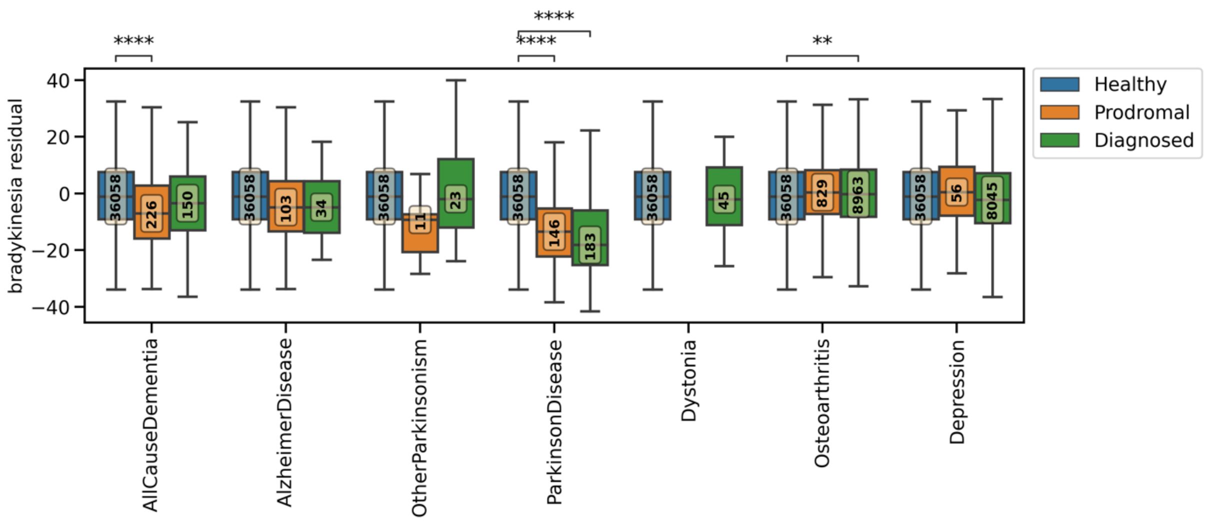


#### Supplemental Figure 23: Most informative feature across models

The group comparisons for the most informative and stable predictor, mean movement during epochs classified as 'light', are shown. The predictor is age, sex, and BMI corrected through coefficients learned from the unaffected control cohort. The boxplot shows the mean and quantiles. The yellow boxes show the number of subjects in each group. The asterisks indicate the significant difference between groups (<0.05 Bonferroni-corrected ns: $3.33x10^{-3} < p <= 1$, *: $3x10^{-4} < p <= 3.33x10^{-3}$, **: $3x10^{-5} < p <= 3x10^{-4}$, ***: $3x10^{-6} < p <= 3x10^{-5}$, ****: $p <= 3x10^{-6}$).

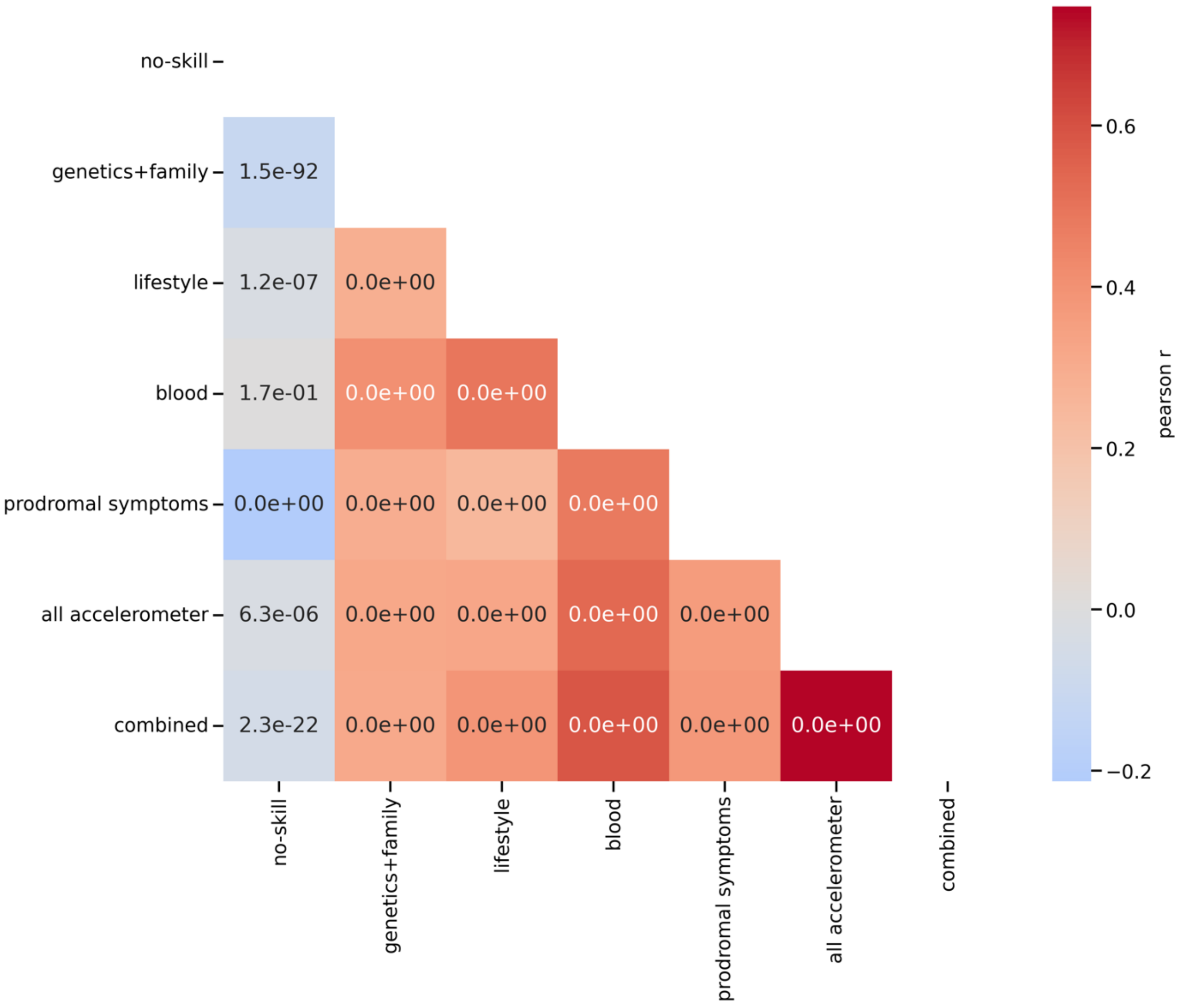


#### Supplemental Figure 24: Correlation of model predictions

The pearson correlation between the mean predicted probabilities of the different models to identify prodromal cases from the population on the test sets is shown. The color indicates the pearson r coefficient with darker red meaning higher correlation and the number indicates the associated p-value. 0.05 Bonferroni corrected significance is reached at 7.14 $x10^{-3}$

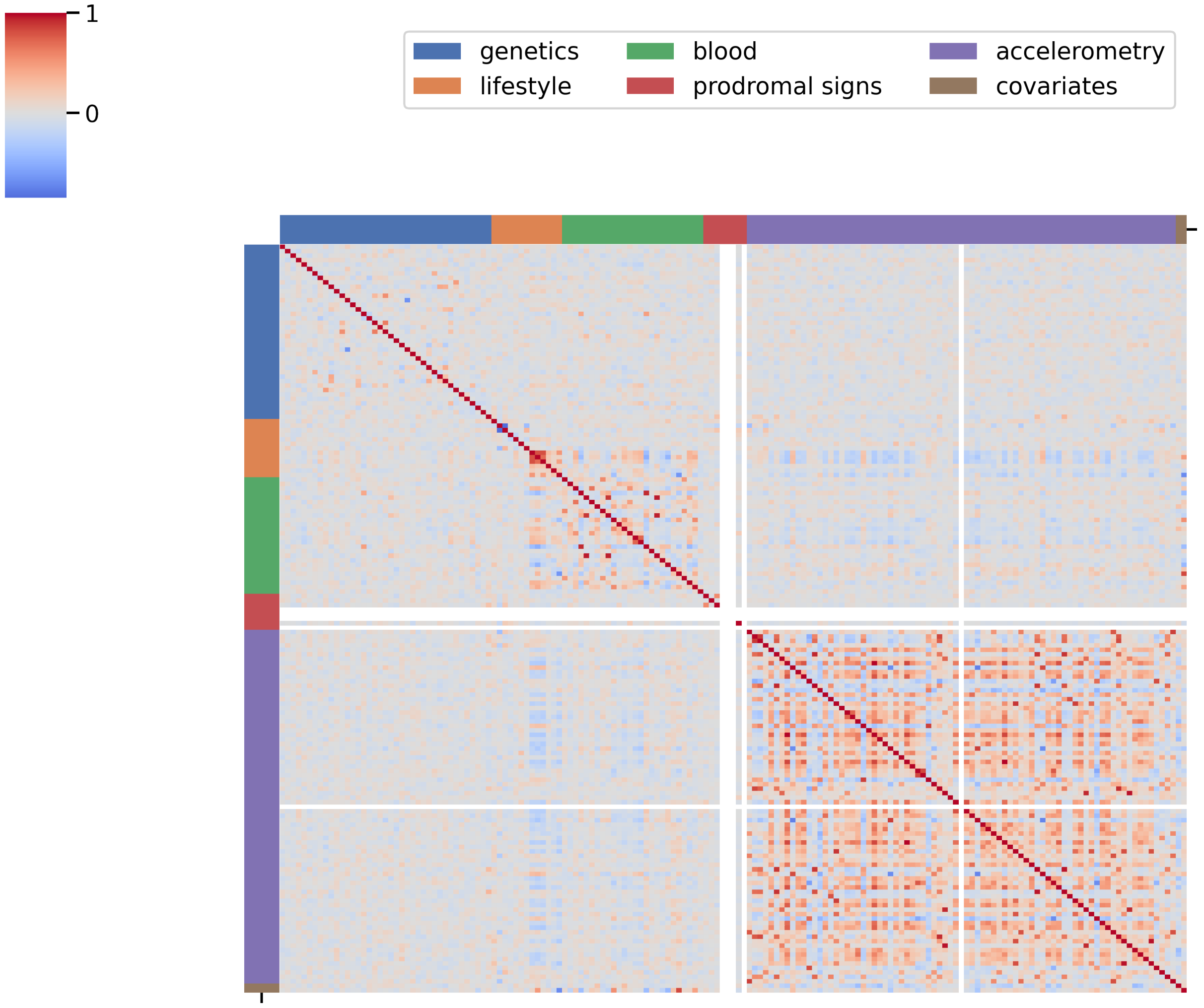


**Supplemental Figure 25: Correlation of predictors**

The pearson correlations between the features of the different modalities in the unaffected controls are shown. The color indicates the pearson r coefficient with darker red meaning higher correlation.

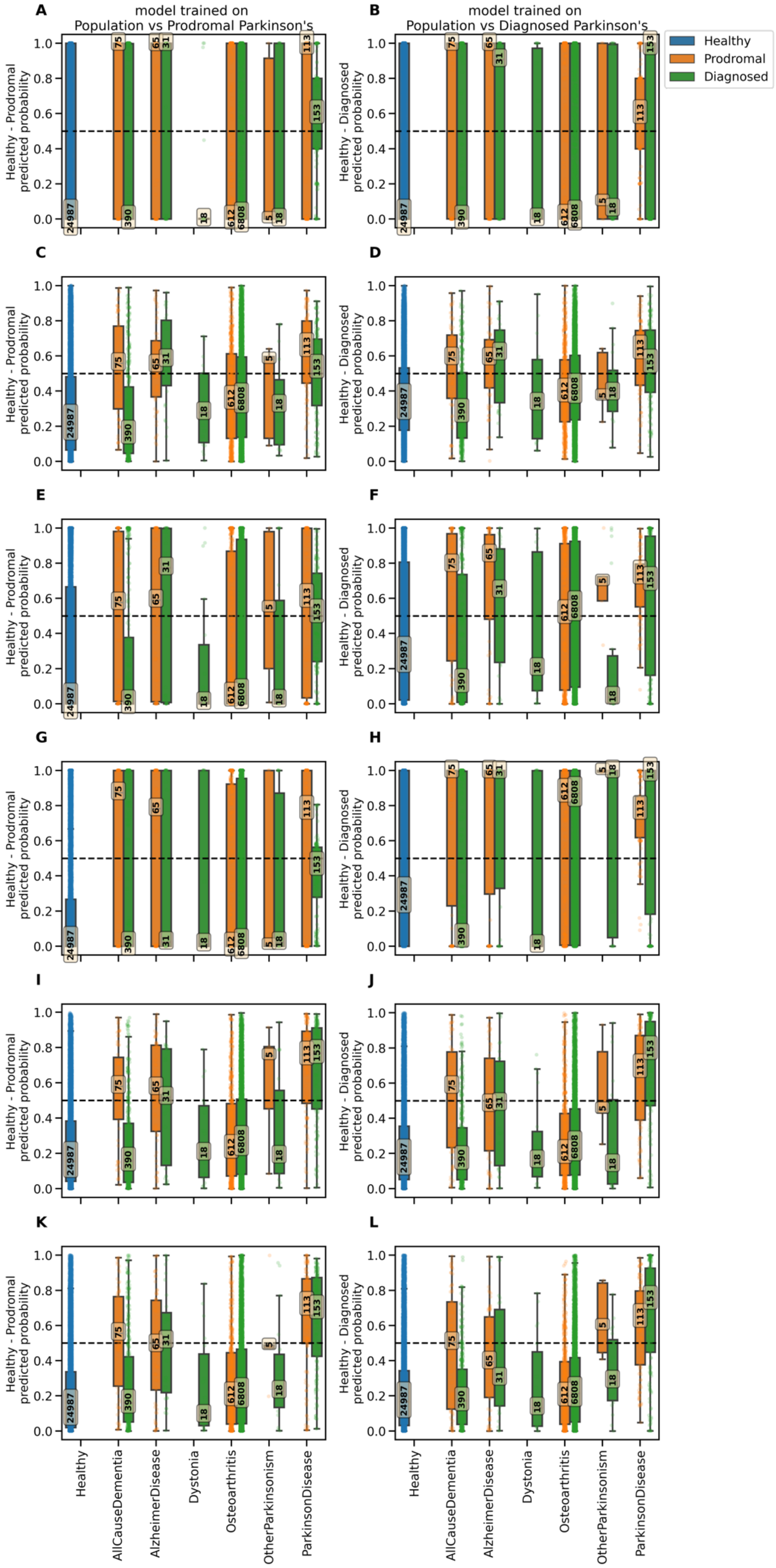

A
model trained on
Population vs Prodromal Parkinson's
B
model trained on
Population vs Diagnosed Parkinson's
Healthy
Prodromal
Diagnosed
C
D
E
F
G
H
I
J
K
L
Healthy - Prodromal
predicted probability
Healthy - Diagnosed
predicted probability
Healthy
AllCauseDementia
AlzheimerDisease
Dystonia
Osteoarthritis
OtherParkinsonism
ParkinsonDisease

#### Supplemental Figure 26: Predicted probability on the test set

The mean predicted probability on the test set across the outer folds is plotted for each diagnosis group. The predicted probabilities are also shown for the external data (not used for training or testing): for the prodromal model, the diagnosed Parkinson's disease (PD) group is used and for the diagnosed model, the prodromal PD group is used. The dashed line indicates a potential 0.5 probability threshold to define the cut-off. This is shown for [left] the prodromal model and [right] the diagnosed model and for each modality-specific model: [A-B] genetics & family, [C-D] blood biochemistry, [E-F] lifestyle, [G-H] prodromal symptoms, [I-J] accelerometry, and [K-L] combined.

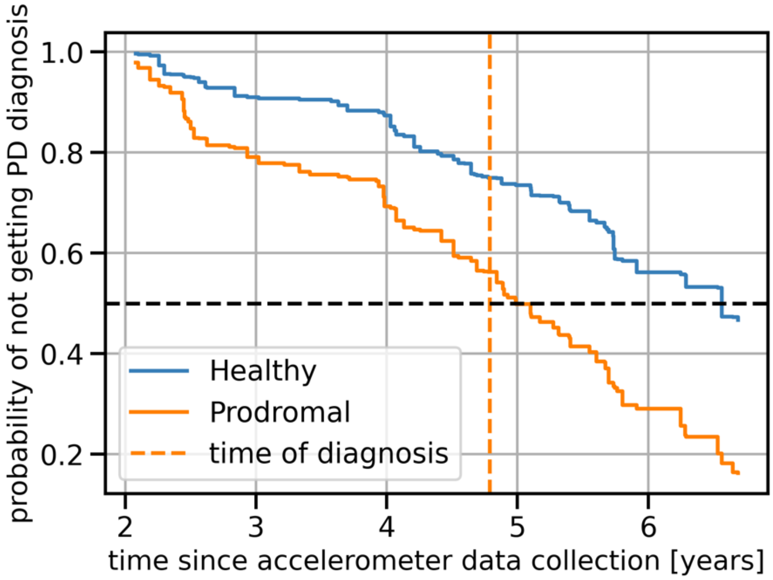


**Supplemental Figure 27: Schematic figure of survival model**

Plot showing the survival function for one Parkinson's disease (PD) prodromal subject and its matched unaffected control. The probability of not getting a diagnosis of PD is shown since the time of accelerometer data collection as estimated by the random survival forest trained on the matched unaffected control setting using all accelerometry features. The intersection of the survival function of the prodromal case with the randomly chosen 0.5 probability threshold (black dashed line) is close to the real time of diagnosis (dashed orange line), meaning that the model correctly predicted the time of diagnosis for this subject.

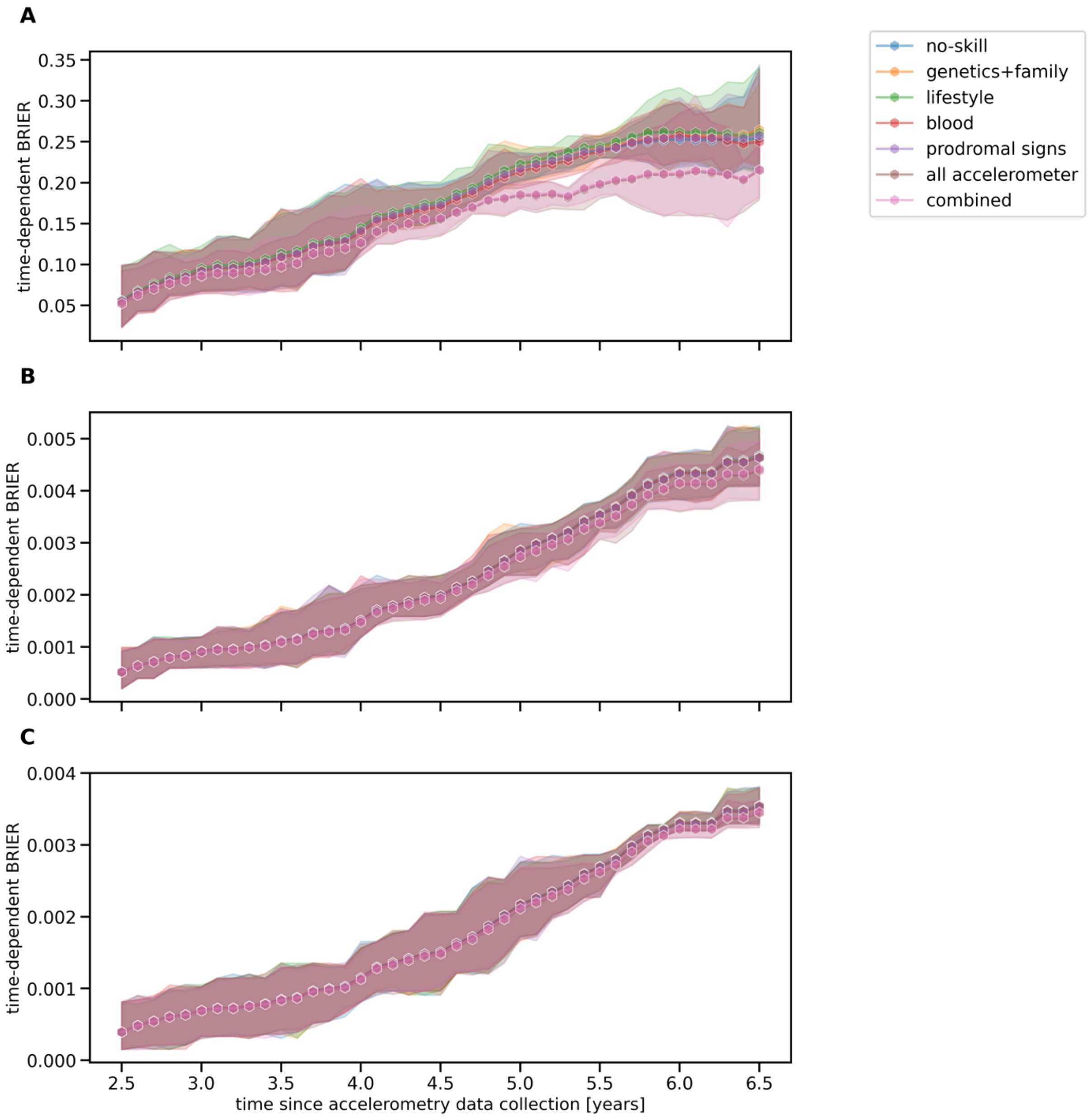


#### Supplemental Figure 28: Brier score for survival models

A performance evaluation of the survival models is provided in a time-dependent manner. The brier score of the random survival forests is plotted for several evaluation time-points (years since data collection) together with Bonferroni-adjusted 95% confidence interval for seven years since accelerometry data collection. We show this for [A] a control group made up of matched unaffected controls, [B] a control group including all unaffected controls, and [C] a control group representing the general population.
